# Differential microRNAs in human serum and sperm after childhood trauma with potential implications for offspring health

**DOI:** 10.1101/2020.08.11.20168393

**Authors:** Ali Jawaid, Magdalena Gomolka, Weronika Tomaszewska, Marina Kunzi, Mahgul Mansoor, Joanna Przybys, Ismail Gbadamosi, Agata Maloburska, Zain Yar Khan, Taufik Hidayat, Anna Chamot, Regina Nadalinska, Muhammad Taha, Adria-Jaume Roura, Bartlomiej Gielniewski, Kristina Thumfart, Safee Ullah Chaudhary, Paulina Fortuna, Łukasz Lewandowski, Mariusz Fleszar, Saba Faisal, Omar Chughtai, Nicola Zamboni, Martyna Plomecka, Susanna Gobbi, Isabelle M Mansuy

## Abstract

Childhood trauma (CT) is associated with prominent psychological and physical effects in humans and may contribute to increased disease susceptibility across generations. Evidence from rodent models suggests that microRNAs (miRNAs) are potential mediators of these effects, including their transmission. We examined serum and sperm miRNAs in three CT human cohorts from a highly consanguineous population via small RNA sequencing and RT-qPCRs. Several miRNAs were differentially expressed in serum of 7-12 years old children with recent paternal loss and maternal separation (PLMS) compared to matched controls. Similar miRNA changes were detected in serum of 18-25 years old male subjects who had been exposed to PLMS in their childhood. Further, some overlapping miRNAs were increased in sperm of adult men exposed to two or more significant traumatic events before the age of 17. Among them, miR-223-3p, a regulator of cholesterol biosynthesis, was consistently upregulated in both serum and sperm across all CT-exposed cohorts. When applied to spermatogonia-like (GC-1) cells in culture, serum from CT-exposed individuals modulated miR-223-3p expression under conditions of altered lipid signaling. Additionally, miRNAs altered in sperm of men with CT were also affected in sperm of mice exposed to early postnatal trauma induced by unpredictable maternal separation combined with unpredictable maternal stress, conditions associated with transgenerational effects. Finally, miR-223-3p mimic delivery into one-cell mouse embryos increased miR-223-3p in the brain and adipose tissue of the resulting animals and caused distinct metabolomic changes in serum. Collectively, these findings suggest that serum and sperm miRNAs may represent molecular correlates of CT, with potential relevance for intergenerational processes and highlight miR-223-3p as a convergent biomarker candidate.

## INTRODUCTION

Exposure to adverse or traumatic experiences during childhood can have enduring consequences for both mental and physical health in humans (Agorastos, 2017; Nelson et al., 2020). Childhood trauma (CT) has been linked to a wide range of adverse outcomes including behavioral impairments, heightened vulnerability to psychiatric disorders, chronic physical illnesses, and metabolic dysfunction in adulthood (Danese & Tan, 2014; Baumeister et al., 2016; Gallo et al., 2017; McKay et al., 2021; Downey & Crummy, 2022; Kuzminskaite et al., 2022; Parvin et al., 2024). Beyond these direct effects, growing evidence suggests that CT is associated with health outcomes in subsequent generations, including the children or grandchildren of exposed individuals. This highlights the importance of understanding how such effects can be carried over generations (Yehuda et al., 2014; Airikka et al., 2023; Dye at al., 2023; Puosi et al., 2024; El-Khalil et al., 2025).

A large body of evidence from rodent models of early postnatal stress demonstrates behavioral deficits and metabolic dysregulation in several offspring generations (Franklin et al., 2010; Gapp et al., 2014; Gapp et al., 2017; van Steenwyk et al., 2018; Boscardin et al., 2022). The biological mechanisms underlying the transmission of phenotypes across generations have become an area of intensive investigations in translational biology. Notably, such heritable phenotypes are thought to be mediated by epigenetic factors, such as changes in DNA methylation and non-coding RNAs, including microRNAs (miRNAs) (Rodgers et al., 2013; Gapp et al., 2014; Rodgers et al., 2015; Gapp et al., 2017; Gapp et al., 2020; Merrill et al., 2021). In mice, overlapping miRNA alterations were observed in brain, serum, and sperm following exposure to unpredictable maternal separation combined with unpredictable maternal stress (MSUS), a model of early postnatal trauma. Further, injecting sperm RNA from adult MSUS males into control 1-cell embryos recapitulated behavioral and metabolic deficits associated with MSUS in the resulting animals (Gapp et al., 2014). These findings highlight a potential role for sperm RNA in the intergenerational effects of early trauma.

In humans, recent studies provide evidence for non-genomic changes in the male germline after exposure to traumatic experiences. Studies have reported DNA methylation and miRNA changes in the sperm of individuals exposed to severe or persistent stressors or trauma during childhood, adolescence or adulthood (Dickson et al., 2018; Roberts et al., 2018; Tuulari et al., 2025). However, no human studies have concurrently examined relevance of non-genomic factors for intergenerational effects across age groups, body fluids, and gametes. Furthermore, only one study conducted comparative analyses of epigenetic/non-genomic signatures of traumatic stress in human cohorts and in ethologically relevant animal models (Dickson et al., 2018).

To address this gap, we assembled three human cohorts with well-documented CT exposure in Pakistan. Pakistani populations were selected due to a higher degree of consanguinity, which may reduce genetic variability within cohorts and facilitate comparison with inbred mouse models (Anwar & Taroni, 2019; Bhinder et al., 2019). We hypothesized that CT is associated with distinct miRNA signatures across biological compartments, including circulation and the germline. We further hypothesized that miRNAs shared across these compartments may be relevant to intergenerational effects. Analyses across the CT human cohorts revealed recent and remote miRNA signatures associated with CT, with some overlap between serum and sperm. Candidate miRNAs altered in human sperm were also examined in sperm of mice exposed to MSUS, a well-characterized model of early postnatal trauma with transgenerational effects. Finally, we experimentally manipulated a convergent candidate, miR-223-3p, in 1-cell mouse embryos as a proof-of-concept approach to assess whether such changes are associated with molecular phenotypes in the resulting animals.

## METHODS

### Human cohorts

This study combined exploratory discovery analyses in three independent human cohorts with targeted validation and mechanistic follow-up. These included a cohort of children with recent paternal loss and maternal separation (PLMS children cohort), adult men with PLMS exposure during childhood (adult PLMS cohort), and men with varying levels of trauma during childhood (CTQ adult cohort). The PLMS children and CTQ adult cohorts served as the primary discovery datasets, whereas the adult PLMS cohort was used for targeted validation of miRNA candidates identified in discovery analyses.

#### 1. PLMS children cohort

To assemble a cohort of children exposed to psychological trauma, we contacted the administration of a residential childcare facility (SOS Children’s Village) in Pakistan and selected children fulfilling the following criteria at the time of assessment: 1) age between 7-12 years, 2) paternal death, 3) maternal separation in the form of adoption by the SOS Village, 4) entry of the child to the SOS Village within 12 months preceding the assessment. Maternal separation was forced because mothers could no longer provide sufficient support to their children and had to transfer their care to the SOS Village. The mothers had no or minimal contact with their children at the time of assessment.

In addition to maternal separation, the children were exposed to deprived parental care in the form of paternal loss. Along with its direct impact on children, paternal loss can also lead to compromised maternal care, as spousal death is a critical life stressor in humans (Moon et al., 2014). Overall, these conditions show similarities to the maternal stress and maternal separation components of the MSUS model (Franklin et al., 2010). Exclusion criteria included 1) history of traumatic brain injury or cerebral palsy, and 2) syndromic or non-syndromic intellectual disability. Based on these criteria, a total of 72 children with paternal loss and maternal separation (PLMS) were initially recruited. A control group comprising 42 children was recruited from the schoolmates of PLMS children. The control group included 7-12-year-old children living with both parents with no history of psychological trauma, traumatic brain injury, intellectual disability or cerebral palsy. Complete confidentiality of participants was maintained through all stages of data collection and analyses. The study procedures were evaluated and approved by the National Head Office of the SOS Children’s Villages, Pakistan. Details of the study were also intimated to the administration of the Global SOS Children’s Villages Organization in Innsbruck, Austria.

##### Demographics

Detailed demographic information for PLMS children was obtained from the administration of the SOS Children’s Village, whereas corresponding information for control children was provided by their parents. This included age, biological sex, and information about the psychological and physical health of the children acquired from their guardians at the time of entry into the SOS Village, as well as reports from the foster mothers. Weight and height were measured in all children by two research interns blinded to the study design. Children were classified as underweight, healthy weight, or overweight based on their correspondence to the ‘less than 5^th^ percentile’, ‘between 5^th^ and 85^th^ percentile’ and ‘85^th^ to 95^th^ percentile’ reference ranges defined for Pakistani children of the same age and sex respectively (Mushtaq et al., 2012).

##### Assessment of depressive and anxiety symptoms

Depressive symptoms in children were evaluated using the Center for Epidemiological Studies Scale for Depression in Children (CES-DC) (Faulstich et al., 1986). CES-DC is a validated tool to screen for depressive symptoms in 6-13-year-old children based on 20 self-report items scored on a Likert-type scale: 0 corresponding to ‘not at all’ and 3 corresponding to ‘a lot’. A score of 15 or higher on this scale indicates a high risk of depression and warrants clinical evaluation and intervention. The anxiety symptoms were evaluated through the Scale for Anxiety Related Disorders (SCARED) (Birmaher et al., 1997). SCARED is a validated 41-item inventory rated on a Likert-type scale: 0 corresponding to ‘never/rarely’ to 3 corresponding to ‘very often’. Two questions related to the parents were not evaluated. CES-DC and SCARED assessments of children were conducted via direct interviews by a research intern blinded to the study design. A confirmation of the children’s responses was obtained from their foster mothers. In case of disagreement between responses (<1% of cases), the responses of the foster mother were considered valid.

##### Serum and saliva collection

Blood was collected by trained phlebotomists blinded to the study design. Blood withdrawal took place during the morning school hours for both groups approximately 1 hour after a standardized breakfast, which followed a period of overnight fasting. All children received a brief explanation of the blood withdrawal procedure and were promised a gift basket for their cooperation. Children showing reluctance or despair were excluded.

After disinfection with a swab, axillary vein venipuncture was performed through a butterfly needle and 6 ml blood was collected per child in serum-separating tube (BD vacutainer, Becton Dickinson). After 1-hour incubation at room temperature, the tubes were centrifuged at 1,300 *g* for 10 min at 4°C for serum separation. Extracted serum was aliquoted into 1.5 ml tubes and stored at −80°C. Saliva was collected by two research interns blinded to the study design. Children who had active upper respiratory tract infections (identified through the symptoms of fever, rhinorrhea, or cough) at the time of sample collection were excluded. All children received a brief explanation of the saliva collection procedure and were promised a gift basket for their cooperation. After a 1-hour period of no oral intake, children were asked to rinse their mouths with clear water twice. Saliva was collected 5 min after rinsing through passive drooling in *salivette* tubes (Sarstedt) over a period of 5 min. Collected saliva was aliquoted into 1.5 ml tubes and stored at −80° C.

#### 2. Adult PLMS cohort

To assemble the adult PLMS cohort, we contacted the administration of the SOS Youth Home in Lahore, Pakistan, which provides lodging facilities for male occupants of the SOS Children’s Village upon turning 15. The criteria for inclusion in the adult PLMS cohort were 1) age between 18-25 years, 2) paternal death and maternal separation in the form of adoption by the SOS Village between the age of 7-12 years. Exclusion criteria included 1) history of traumatic brain injury, intellectual disability or cerebral palsy, 2) history of drug abuse. Based on these criteria, a total of 13 adults with paternal loss and maternal separation (PLMS) were selected. A control group (n=16) was recruited from 18-25-year-old college students from the same vicinity who lived with both parents at least till the age of 18 and with no history of psychological trauma, traumatic brain injury, intellectual disability, cerebral palsy, or drug abuse. Complete confidentiality of participants was maintained at all stages of data collection and analyses. The study procedures were approved by the National Head Office of the SOS Children’s Villages, Pakistan.

#### 3. CTQ adult cohort

The semen samples from adult men were collected in collaboration with Chughtai Health, Pakistan, a leading diagnostic and outpatient clinical care facility in the country. Patients visiting the lab for routine seminal analyses were invited to participate in the study. Those who consented were provided a free-of-charge assessment of their reproductive health. After consenting and routine semen collection procedures, participants were assessed for CT exposure using a structured self-report questionnaire adapted for the Environmental Influences on Child Health Outcomes (ECHO) cohort (Version 01.20, November 30, 2018). Although referred to in the original documentation as the ‘Childhood Trauma Questionnaire (CTQ)’, this instrument does not correspond to the CTQ developed by Bernstein et al. (2014) but instead constitutes a harmonized trauma exposure measure informed by the original CTQ framework. This questionnaire addresses six traumatic experiences prior to the age of 18 (parental death, parental divorce, violence, sexual abuse, illness or other) and the perceived severity of the event. Basic demographic and clinical information were also collected from the participants by a research intern blinded to the study design. Personal information was coded, and the data was handled with discretion and anonymity. The collected samples were analyzed through automated procedures. The assessment included appearance, consistency, pH, % of normal cells, % of motile cells, and progression stages. After analysis, the semen samples were centrifuged at 500 *g* for 5 min at 4 °C to pellet out the sperm. The sperm pellet and the seminal fluid were stored separately at −80 °C.

### Sample size considerations for human cohorts

A priori sample size considerations were used during then planning stage to guide recruitment of the human cohorts with an aim of n=50 CT-exposed subjects and controls each in both discovery cohorts. Because the study involved biofluid collection in vulnerable and constrained populations, as well as downstream exclusion of samples, the final analytic sample sizes differed across assays. Accordingly, we distinguish between planned cohort recruitment and achieved sample sizes used for specific discovery and validation analyses.

The PLMS children cohort initially included 72 children along with 42 controls. However, blood and psychological profiling was fully completed for n=70 PLMS and n=35 control children. Similarly, 48 PLMS and 18 control serum samples were available for small RNA sequencing, and only 22 PLMS and 8 control serum samples were available for targeted RT-qPCR validation (Extended Data Fig. 1A). The adult PLMS cohort comprised 13 PLMS and 16 control men and was designed as a targeted validation cohort rather than an unbiased discovery cohort (Extended Data Fig. 1B). The adult CTQ cohort comprised 93 men in total. Among these, sperm samples from 26 individuals with no trauma (labelled CTQ0), 17 individuals with single (labelled CTQ1), and 23 with two or more traumatic events (labelled CTQ2) were included in the small RNA sequencing. Subsequently, 26 CTQ0, 14 CTQ1 and 22 CTQ2 samples were used for the main RT-qPCR validation analyses (Extended Data Fig. 1C).

For the primary discovery analyses based on group-wise comparisons, the achieved sample sizes in the PLMS children serum cohort (n=48 PLMS vs n=18 control children) and adult CTQ cohort (n=23 CTQ2 vs n=26 CTQ0) were considered sufficient to detect biologically meaningful moderate-to-large between-group differences, approximately in the range of 30-50%, using two-sided testing with α set at 0.05. By contrast, the smaller adult PLMS cohort (n=13 adult PLMS vs n=16 adult control) was not considered sufficiently powered for unbiased discovery and was therefore used only for targeted validation of candidate miRNAs identified in the discovery cohorts. Given the exploratory nature of the sequencing analyses and the limited prior literature available to estimate expected CT-associated miRNA effect sizes in human serum and sperm, discovery findings were interpreted as hypothesis-generating and followed by targeted RT-qPCR confirmations wherever sample material was available.

### Cell culture and siRNA-mediated knockdown

Commercially available GC-1 spermatogonia-like cells (ATCC, CRL-2053) were cultured at 37 °C with 5% CO_2_ in a biosafety level 2 (BSL2) cell culture laboratory. Culture medium was composed of high-glucose DMEM with 10% fetal bovine serum (FBS, HyClone) and 40 μg/ml gentamicin (Sigma Aldrich). GC-1 cells have characteristics of a stage between type B spermatogonia and primary spermatocytes. For knockdown of *SCARB1* using RNA interference, approximately 3000 cells per well were plated in 500 μl culture medium in 24-well plates and incubated overnight. Protocol per well: Before transfection, the medium was removed, the cells washed, and 400 μl antibiotic free medium added. 1 μl siRNA in 49 μl OptiMEM and 1 μl Lipofectamine 2000 (Invitrogen) in 49 μl OptiMEM were incubated separately for 5 min and subsequently mixed and incubated for 20 min to allow formation of transfection complexes. 100 μl of transfection mixture was added dropwise to each well. siRNAs used were siRNAs targeting *SCARB1* (Flexitube siRNA, Qiagen) or negative control siRNA (All Star negative control, Qiagen). 24 h after siRNA transfection, the cells were treated with 10% pooled serum from the adult PLMS and control cohorts respectively for 24 h. The cells were then harvested by removing the medium, washed with ice-cold PBS and lysed with QIAzol reagent (Qiagen) for RNA extraction.

### MSUS mice

C57BL/6J mice were housed in a temperature- and humidity-controlled environment under a reverse light-dark cycle, with unrestricted access to food and water. All experimental procedures were carried out during the animals’ active phase, following the guidelines and regulations of the Cantonal Veterinary Office in Zurich.

To model unpredictable maternal separation combined with unpredictable maternal stress (MSUS), C57BL/6J dams (aged 2–3 months) and their litters were randomly assigned to the experimental group. From postnatal day (PND) 1 to 14, pups were subjected to daily 3-hour separations from the dams during which the mothers were exposed to acute stress in the form of cold swim or restraint in an unpredictable fashion (Franklin et al., 2010). Control litters were left undisturbed except for a weekly cage change until weaning at PND 21. After weaning, male offspring were housed in social groups of 4–5 mice per cage, grouped by treatment but from different litters to minimize litter-specific effects.

Mouse sperm was collected using the swim-up method as described previously (Gapp et al., 2014) with minor modifications. Briefly, the cauda epididymis was dissected, placed in 1500 µl M2 medium (Sigma-Aldrich) in 2 ml tubes for 20 minutes to 1 hour, perforated with dissection scissors to allow mature sperm to swim out of the tissue. Medium was then incubated at 37 °C for 1 hour to allow mature sperm to swim up. Next, the upper 500 µl of the medium was filtered, washed with ice-cold PBS and centrifuged at 2,600 *g* for 6 min at 4 °C. Supernatant was discarded and sperm pellet was resuspended in 400 µl ice-cold PBS. Sperm solutions were stored at –80 °C until further use.

Prior to sacrifice, mice were fasted for 6 hours during the second half of the inactive phase. Next, mice were deeply sedated with 5% isoflurane in inhaled air and euthanized by cervical dislocation. Death was confirmed by decapitation; whole blood was collected from the site of decapitation into non-coated Eppendorf tubes and immediately put on ice. For clotting, blood was left at room temperature for 30-60 min, centrifuged at 2,000 *g* for 10 min at 4 °C. The supernatant (serum) was collected and stored at −80 °C.

### Embryo modification using miR-223-3p mimic

Superovulation was induced by hormonal stimulation of C57BL/6J females (5-6 weeks old) using pregnant mare serum gonadotropin (PMSG) followed by human chorionic gonadotropin (hCG) after an interval of 48 hours. Females were mated with males immediately after hCG administration.

The next morning, females with visible seminal plug (0.5 days post coitum) were sacrificed by cervical dislocation and oviducts were collected. At this stage, embryos are naturally present at the ampulla of oviduct, which is the widest section of the fallopian tube where fertilization occurs. By disrupting the wall of the ampulla, 1-cell stage embryos were collected and incubated in pre-warmed M2 medium with hyaluronidase (300 µg/ml) for 5 min to release attached cumulus cells. Embryos were then picked using a glass capillary connected to a mouth-controlled aspirator. Embryos were subsequently washed in 60 mm petri dish containing several drops of M2 medium covered with liquid paraffin to remove cellular debris and then transferred into a new plate with 3 drops (100 µl) of KSOM medium (pre-equilibrated in an incubator at least 15 min prior to isolation).

miRNA mimics were introduced into the embryos through electroporation using a BioRad Gene Pulser Xcell electroporator. The following mimics were introduced: a) mmu-miR-223-3p miRCURY LNA miRNA mimic (Qiagen, 339173/ YM00471011-ADA), b) Negative Control 4 (NC) miRCURY LNA miRNA mimic (Qiagen, 339173/ YM00479903-ADB). Electroporation mixtures were prepared according to the formulation 0.25 ml OptiMEM + 50 embryos + 1 µl of 1 µM miRNA mimic/control (final concentration: 4 nM). The mix was incubated at RT for 10 min followed by electroporation performed under the following parameters: Voltage (V): 30, pulse length (ms): 2, number of pulses: 2, pulse interval (s): 0.1, cuvette (mm): 1. After electroporation, embryos were incubated (37 °C, 5% CO_2_) overnight in KSOM medium. Mimics were labelled with FAM, a fluorescent dye emitting strong green signal when excited by a specific wavelength (∼494 nm), which allowed observation under fluorescent microscope.

One day before embryo transfer, CD1 females were mated with vasectomized males to induce pseudopregnancy. Females with visible seminal plugs (0.5 days post coitum) were used as surrogates (embryo recipient). For embryo transfer procedures, designated surrogates were weighed and anesthetized via IP injection of ketamine (50-75 mg/kg) and medetomidine (0.5-1 mg/kg) in saline solution. This was followed by subcutaneous injections of butorphanol (3.3 mg/kg), tolphedine (2 mg/kg), and enrofloxacin (2.5 mg/kg) for their analgesic, anti-inflammatory, and antibiotic properties. During the procedure, eyes were covered with ointment (Vita Pos) to prevent corneal drying. Fur was removed on the back, and skin was disinfected with ethanol and iodine. Additional local anesthetic (100 µl of 0.25% bupivacaine) was injected at the incision site. A small incision was made with fine scissors at the dorsal midline (below the last rib), perpendicular to the vertebral column, to allow access to both oviducts. Subsequently, the ovarian fat pad was grasped with blunt forceps, and the ovary, oviduct and uterus were pulled out of the body cavity and placed on the sterile gauze. Reproductive tract was immobilized on the gauze using serrefine clamp clipped on the ovarian fat pad. Ampulla of the oviduct was localized under a stereomicroscope. A transfer pipette loaded with embryos (∼13) was inserted into the oviduct through the small incision in oviduct wall or through the infundibulum after breaking the bursa, a transparent membrane surrounding the ovary and oviduct. Embryos were loaded into the glass pipette in the smallest possible volume of M2 medium between air bubbles. Proper transfer was confirmed by observing the air bubbles inside the oviduct. The reproductive system was gently inserted back into the abdominal cavity, and the procedure was repeated on the other oviduct. The body wall was then sutured using Safil surgical suture (4/0 needle 26 mm 1/2 c; absorbable suture made of polyglycolic acid), while the skin was closed using wound clips (9 mm). To shorten anesthetic effects Atipam (0.5-1 mg/kg) was injected (IP) as well as a saline (0.5 ml; SC.) to prevent dehydration. The animal was placed in the clean cage on the warming plate until full recovery from anesthesia.

The pregnancy lasts ∼21 days in CD1 mice. The offspring after birth were left unperturbed with the surrogate CD1 mothers, weaned at day 21, and maintained under standard housing conditions until the age of 1 year. Prior to sacrifice mice were fasted for 6 hours during the second half of the inactive phase. Next, mice were deeply sedated with 5% isoflurane in inhaled air and euthanized by cervical dislocation. Death was confirmed by decapitation; whole blood was collected from the site of decapitation into non-coated Eppendorf tubes and immediately put on ice. For clotting, blood was left at room temperature for 30-60 min, centrifuged at 2,000 *g* for 10 min at 4 °C, the supernatant (serum) was collected and stored at –80 °C. Following decapitation, the brain, white adipose tissue (WAT), and brown adipose tissue (BAT) were rapidly dissected, snap-frozen in RNAlater stabilizing solution (Invitrogen) in dry ice and stored at –80 °C.

### RNA extraction and real-time quantitative PCR (RT-qPCR)

#### Human samples

RNA extractions from human serum, sperm, and seminal fluid were performed using QIAzol (Qiagen) or TRIzol (Invitrogen) reagents. Briefly, 1 ml QIAzol/TRIzol was added to 200 µl of serum, 200 µl of seminal fluid or to the sperm pellet, respectively. This was followed by chloroform addition, aqueous phase separation, and isopropanol-based precipitation of RNA. The extracted RNA was quantified using NanoDrop spectrophotometer and reverse-transcribed using miRCURY LNA RT Kit (Qiagen). RT-qPCR was performed on the resulting cDNA according to miRCURY LNA SYBR Green PCR Kit (Qiagen) using the StepOnePlus™ Real-Time PCR System. Selected miRNAs were amplified by specific miRCURY LNA miRNA primers for hsa-miR-223-3p, hsa-miR-144-3p, hsa-miR-145-5p, hsa-let-7b-5p, hsa-miR-29a-3p, hsa-miR-30b-5p, hsa-miR-25-3p, hsa-miR-141-3p, hsa-miR-21-5p, hsa-miR-29c-3p, hsa-miR-148a-3p, hsa-miR-101-3p, hsa-miR-320e, and miR-449a-5p. Small nuclear RNA (RNU6) or spike-in control (UniSP6) were used as controls for relative expression analyses in sperm and serum respectively.

For quantification of miR-16-5p, miR-34c-5p, miR-375-5p, and miR-449-5p in human sperm, extracted RNA was reverse transcribed using miScript II RT kit (Qiagen). Subsequently, RT-qPCR was performed with miScript primer assays (Qiagen) on LightCycler 480® (Roche) for hsa-miR-16-5p, hsa-miR-34c, hsa-miR-375-5p, and hsa-miR-449a-5p. Small nuclear RNA (RNU6) was used as endogenous control for relative expression analyses.

#### Mouse samples

RNA extraction, reverse transcription, and RT-qPCR from mouse serum were performed as described for human samples without any major modifications. RNA extractions from mouse sperm, brain, WAT, and BAT were performed following the same protocol with addition of an initial homogenization step of the tissue before adding Qiazol. Extracted RNA was reverse transcribed using miRCURY LNA RT Kit (Qiagen). RT-qPCR was performed as described above using RNU6 and UniSp6 as controls for relative expression analyses in sperm and serum respectively.

#### GC-1 spermatogonia-like cells

For RNA extraction, the medium was removed from the plate, and the cells were washed three times with ice-cold PBS, lysed and homogenized by adding 1 ml of TRIzol reagent (Invitrogen) directly to the plates. After 5 min of incubation, followed by chloroform addition, aqueous phase separation, and isopropanol-based precipitation of RNA. After quality assessment, extracted RNA was reverse transcribed using miRCURY LNA RT Kit (Qiagen) and RT-qPCR was performed as described above.

The sequences for the Qiagen primers used for all RT-qPCR assays are collectively listed below:

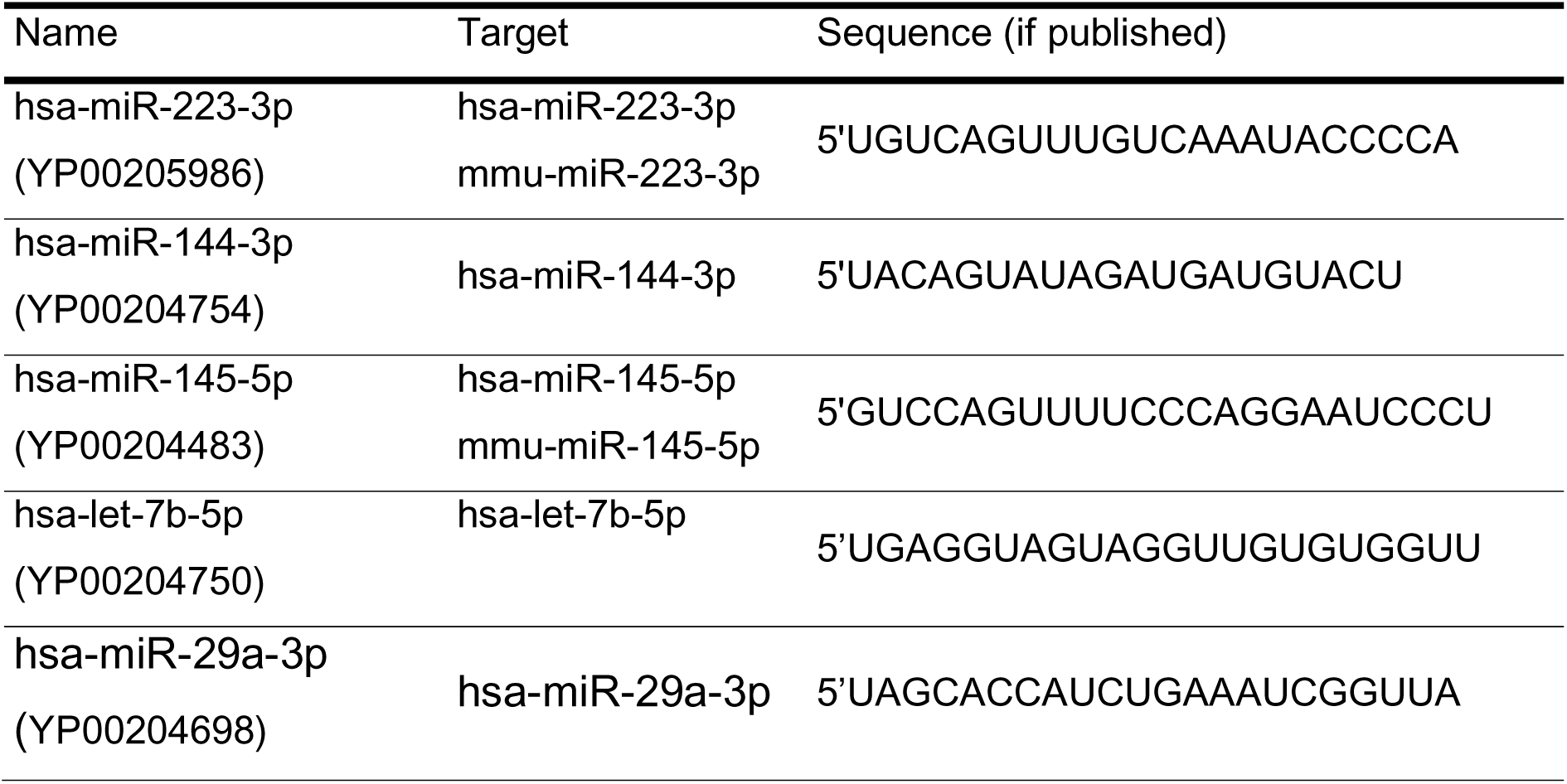
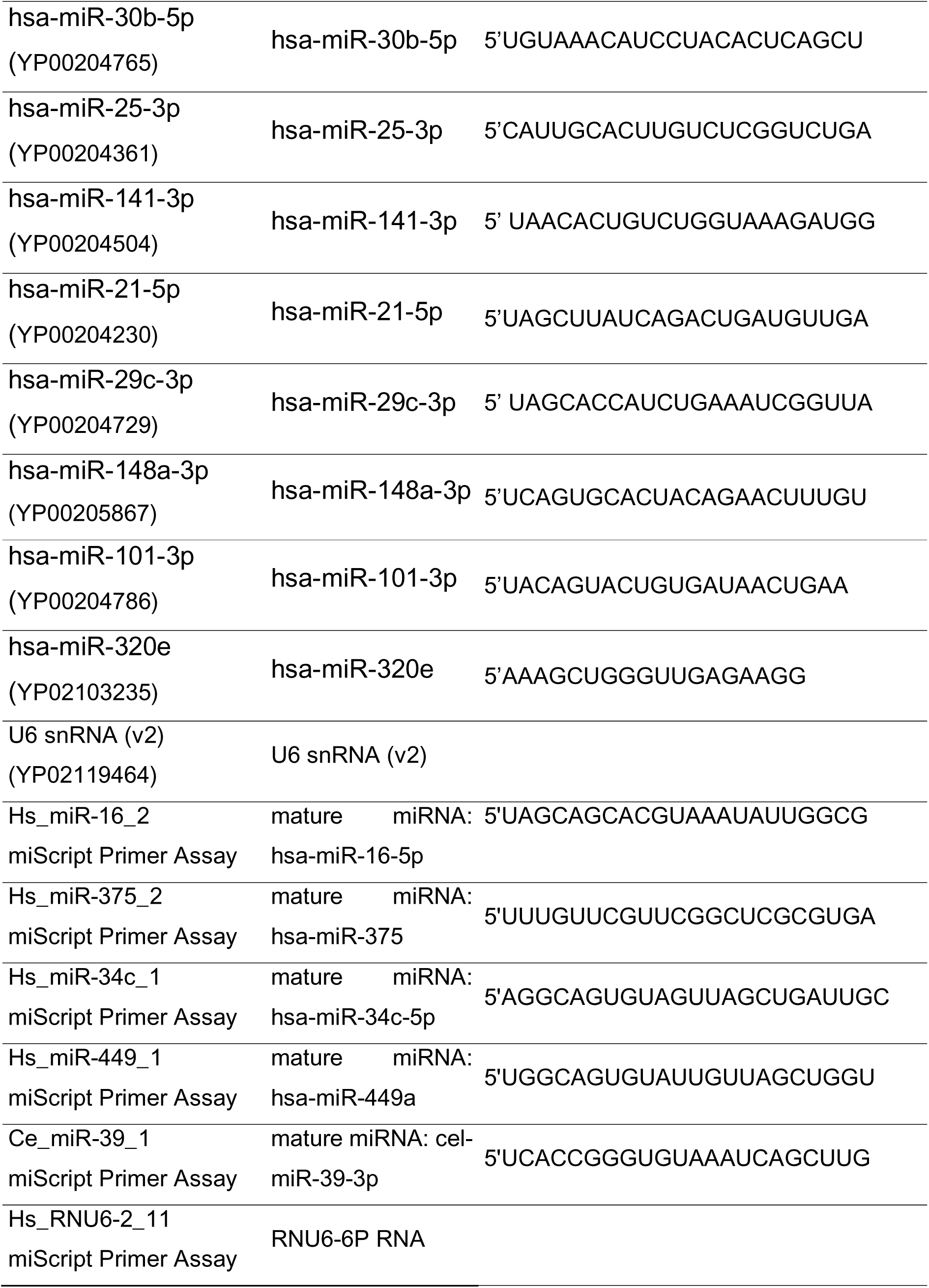

### Small RNA sequencing including statistical analysis

Quality and integrity of the isolated RNA were assessed using an Agilent 2100 Bioanalyzer with an RNA 6000 Nano Kit (Agilent Technologies). miRNA libraries were prepared using the QIAseq miRNA Library Kit according to the manufacturer’s protocol (Qiagen). Briefly, miRNA libraries were generated from 200 ng of total RNA. The initial step involved ligation of specific adapters to the 3′ and 5′ ends, enabling universal reverse transcription. cDNA was then synthesized by reverse transcription and purified using specialized purification beads provided by the manufacturer. The purified cDNA was subsequently amplified by PCR to obtain sufficient miRNA library concentration. Library evaluation was performed with the Agilent 2100 Bioanalyzer using the Agilent DNA High Sensitivity chip (Agilent Technologies). The mean library size was 174 bp. Libraries were quantified using a Quantus fluorometer and the QuantiFluor double-stranded DNA System (Promega). Finally, libraries were single-end sequenced (75 bp) on the NovaSeq 6000 platform (Illumina).

Qiagen GeneGlobe Data Analysis Center (RRID:SCR 021211) was used for primary miRNA bioinformatic analysis. The experiment’s unique molecular identifiers (UMIs) were quantified in five steps: 1) calibration of miRBase entries, 2) trimming of adapter and low-quality bases, 3) identification of UMIs, 4) sequence alignment using the MiRBase V21 and piRNABank databases, and 5) quantification of unique molecules. The resulting primary miRNA quantification can be found in the *QIAGEN_primary_analysis.summary.odf* file. Samples with miRNA read counts less than 200,000 were excluded from further analysis.

For the secondary bioinformatics analysis, a DESeq2 object was built with UMIs and associated clinical data. Non-expressed or low-expressed miRNAs were discarded during the pre-filtering step. Because the studied tissues (sperm and serum) differed significantly, they were examined separately, and miRNA expression differences between controls and the studied groups were determined (PLMS vs control children; CTQ0 vs CTQ2 adults). In the serum dataset, biological sex of the subjects was used as a covariate in the differentially expressed analysis formula. To estimate the normalized differentially expressed miRNA levels across groups of interest, the plotCounts function was used. The EnhancedVolcano package (https://github.com/kevinblighe/EnhancedVolcano) was used to better visualize miRNA differences between groups.

### Metabolomic assessments including statistical analysis

Samples for serum metabolomic analysis were precipitated using ice cold methanol in 1:4 serum: solvent ratio. The entire mixtures were then transferred to a microcentrifuge tube, vortexed for 10 min at 4 °C and frozen at −80 °C for 1 hour. Next, the tubes were centrifuged at 12,000 *g* at 4 °C for 7 min. 350 µl of the supernatant from each sample was collected for metabolomics analysis and dried using a centrifugal vacuum concentrator (HETOVAC) at 35 °C. The resulting precipitate was re-dissolved in 50 µl of 3% methanol in water and transferred to autosampler glass vials for liquid chromatography-mass spectrometry (LC-MS) analysis.

Separation of analytes was performed with ACQUITY ultra-high performance liquid chromatography (UHPLC) system using a normal-phase (Amide) column with a gradient elution of solvent A (water containing 0.1% formic acid) and solvent B (acetonitrile containing 0.1% formic acid). Analytes were separated using ACQUITY UPLC BEH Amide (1×100 mm,1.75 µm) with a linear gradient from 95% to 30% of mobile phase B in 25.5 min with a total flow rate of 180 µl/min.

Mass spectrometry was performed using a Synapt XS ion-mobility quadrupole-time of flight mass spectrometer operated in positive and negative polarity mode. Data was acquired in data-independent acquisition (DIA) mode using the MSE data acquisition function. In function one, a low collision energy of 6 eV was applied, while in function two, a high collision energy was ramped from 10 to 35 eV for fragmentation. Leucine-Enkephalin was used as a lock mass (m/z 555.2771 for positive ionization and m/z 554.2615 for negative ionization). The source parameters were set as follows: nebulizing gas (nitrogen) at 800 L/h, drying gas at 65 L/h, spray voltage at 2.8 kV, source temperature at 120 °C, and desolvation temperature at 350 °C.

The MS data were processed using Progenesis QI (Nonlinear Dynamics, UK version 3.0) and MS-DIAL (version 5.2) from RIKEN Center for Sustainable Resource Science: Metabolome Informatics Research Team to perform spectral deconvolution, peak identification, alignment, normalization, and compound identification against the Human Metabolome Structure Database. The resulting data were then subjected to statistical analysis using the web-based tool MetaboAnalyst 6.0 (https://www.metaboanalyst.ca). Raw metabolite intensity measurements were initially grouped by metabolite identifiers, with intensities summed across all entries sharing the same ID, regardless of ionization adduct or ontological classification. This procedure removed technical redundancy and streamlined downstream analyses.

Sample identifiers were parsed into unique components, including sample ID, replicate number, and experimental group. For each metabolite, technical replicates within a biological sample were averaged to yield a single representative intensity value. In parallel, metabolite intensities were aggregated at the ontology level by summing values across all metabolites sharing the same classification. These class-level intensities were then averaged across replicates within each sample. The resulting datasets - one at the individual metabolite level and one aggregated by ontology - were merged by sample ID and experimental group into a unified feature matrix. This approach preserved both fine-grained metabolite data and broader functional summaries, facilitating comprehensive analysis.

For descriptive visualization, data are reported as means ± 95% confidence intervals (CI), calculated using the t-distribution. Between-group comparisons were conducted using the nonparametric Mann–Whitney U test, chosen for its robustness to non-normal distributions and outliers. Due to the exploratory nature of the study, no correction for multiple testing was applied, and thus results should be interpreted as hypothesis-generating rather than conclusive.

To enable comparison across chemically diverse metabolite classes, intensity values were scaled to the [0,1] range using Min-Max normalization. This method preserved relative differences while minimizing distortions caused by variations in magnitude typical of omics data. The normalized values were used to generate radar plots illustrating metabolite ontology profiles across experimental groups, facilitating visual comparison of metabolic shifts. All data processing and visualization steps were performed using Python (version 3.13.3), employing core packages including *pandas*, *numpy*, *scikit-learn*, and *matplotlib*.

### Statistical analyses of behavioral and molecular data

Statistical analyses of the human psychological data, blood parameters, and qPCR assays, as well as MSUS sperm RT-qPCRs were performed with GraphPad Prism version 10 (GraphPad Software). Outliers were removed using the ROUT test (Q=1%), and the fold changes were adjusted to the mean of the respective control group, wherever possible. The datasets were then checked for normality with the Shapiro-Wilk test. Normally distributed data was analyzed with an unpaired parametric t-test (comparison of two groups) or one-way ANOVA (comparison of more than 2 groups) with Tukey’s post hoc correction. Data from CES-DC was exceptionally analyzed by unpaired t-test despite non-normal distribution considering the ordinal nature of the data. However, Welch’s correction was performed to account for differential variances of the two groups. Remaining non-normally distributed data was analyzed with the Mann-Whitney U non-parametric test (comparison of two groups) or the Kruskal-Wallis test (comparison of more than 2 groups) with Dunn’s post hoc correction. The effect of SR-B1 knockdown on the expression of miR-223-3p in GC-1 cells treated with control vs adult PLMS serum was examined through two-way ANOVA. A *p* value <0.05 was considered statistically significant for comparisons, whereas *p* value >0.05 but <0.1 was considered a trend towards significant difference. All graphs were drawn with GraphPad Prism version 10, with representation of individual data points, wherever applicable.

### Ethical considerations

Informed consent was obtained from all adult participants for anonymous analyses of their serum, sperm, and seminal fluid samples. For the PLMS children’s cohort, the administration of the SOS Village authorized the consent, whereas the consent for data and sample collection from control children was provided by their parents. The confidentiality of the participants was maintained throughout all phases of the study. The study procedures were reviewed and approved by the National Office of the SOS Children’s Villages, Pakistan and the executive board of Chughtai Healthcare, Pakistan. Experiments with mice were run in conformity with guidelines of the Cantonal Veterinary Office of Zurich and the Swiss Animal Welfare Act (Tierschutzgesetz) under the license number ZH021/2022. Animals were maintained in a reverse cycle and were tested during their active cycle (darkness).

### Data and code availability

The serum and sperm RNA sequencing data from the study have been placed on open repositories. Analysis code is available at GitHub: (https://github.com/gomolkamagda-lena1/miRNAseq_PLMSandCTQ_analysis). Raw sequencing data and processed count files are available in ArrayExpress/BioStudies (accession number listed in the readme file available on https://github.com/gomolkamagdalena1/miRNAseq_PLMSandCTQ_analysis). A separate file containing supplementary statistics for all the main and supplementary figures is available as part of the submitted *Supplementary Material*. Further data are available from the corresponding authors upon request for research purposes only and subject to institutional and ethical restrictions. The bioinformatic and statistical analyses were performed using openly available software packages as specified in the Methods section. Custom analysis scripts are available from the corresponding authors upon reasonable request.

## RESULTS

### Psychological symptoms and altered blood parameters in children with CT exposure

The first cohort comprised 7-12 years-old children (n=72) residing in SOS Children’s Village, Pakistan. SOS Children’s Village operates as a residential childcare facility that provides long-term foster care to children who have been separated from their parents. The children included in the cohort were exposed to paternal loss and maternal separation (PLMS) within the preceding year. Paternal loss was selected as an exposure because spousal death represents a major stressor for mothers who, in this context, were compelled to place their children in the SOS Children’s Village because of limited financial resources (Prior et al., 2018). This sequence of maternal stress followed by maternal separation partially recapitulates the maternal stress and separation components of the MSUS mouse model of postnatal trauma (Franklin et al., 2010; Gapp et al., 2014). A group of control children (n=42) living with both parents was recruited from the schoolmates of PLMS children. PLMS and control children were matched for age, gender and body mass index (BMI). BMI was included as a descriptive anthropometric measure because more detailed long-term metabolic assessments were not available. Further, both groups had comparable hemoglobin levels, providing indirect information about micronutrients such as iron, folate, and vitamin B12 (Supplementary Table 1).

Subsequent blood and psychological assessments were fully completed for n=70 PLMS and n=35 control children, and the results therefore reflect these subsets only. Both groups underwent thorough behavioral assessment by blinded investigators using the Center for Epidemiological Studies Depression Scale for Children (CES-DC) (Faulstich et al., 1986) and the Scale for Anxiety-Related Disorders (SCARED) (Birmaher et al., 1997) in children. The results showed significantly higher scores (*p<0.05*) on the depression scale in PLMS children (15.27 ± 7.25) compared to controls (12.63 ± 2.95) (Fig. 1A). Additionally, the PLMS group has a higher percentage (*p<0.05*) of children that could be classified as depressed based on the CES-DC cut-off for depression (Faulstich et al., 1986, Fig. 1B). However, a similar pattern was not observed for anxiety disorders (Supplementary Fig. 1). These behavioral manifestations were accompanied by an increased erythrocyte sedimentation rate in children with PLMS (median [IQR]: 8 [5–12.5] vs 7 [5–8] mm/h in controls; *p<0.05*; Fig. 1C). PLMS children also had higher serum triglyceride (TG) levels (median [IQR]: 106 [76.5–145.3] vs 79 [60–100] ng/ml in controls; *p < 0.01*) but only non-significant changes in the other serum lipids, including high-density lipoproteins (HDLs) and low-density lipoproteins (LDLs, Fig. 1D). To aid clinical interpretation, serum lipid measures were additionally classified into desirable and undesirable ranges using established pediatric reference cut-offs based on the *Integrated Guidelines for Cardiovascular Health and Risk Reduction in Children and Adolescents* by the National Institute of Health, USA (National Heart, Lung, and Blood Institute, 2012) and the American Heart Association (Peterson et al., 2026). The proportion of children with TG levels in the non-desirable range was greater in the PLMS group than in controls (34% vs 11%; *p<0.05*; Extended Data Fig. 2). Finally, unbiased metabolomic analysis of serum from PLMS and control children revealed an enrichment for metabolites associated with glycerophospholipid, glycerolipid, and taurine and hypotaurine metabolism in the serum of PLMS children (Fig. 1E).

**Figure 1.**
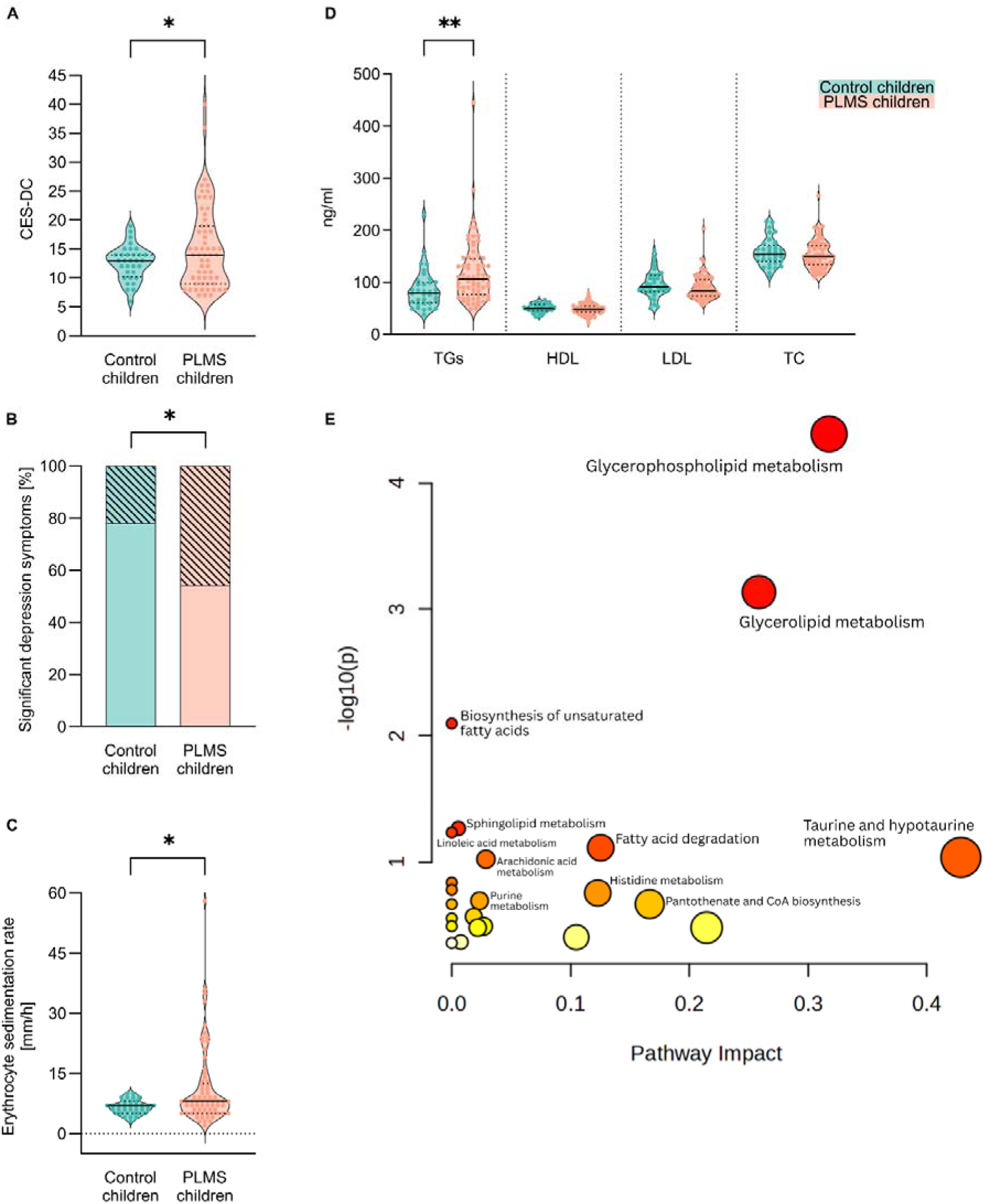
Psychological symptoms and blood parameters in PLMS and control children. **(A)** CES-DC (depression) scores in PLMS (n=59) and control children (n=32). Mean ± SD: 15.27 ± 7.25 PLMS vs 12.63 ± 2.95 control; unpaired t-test with Welch’s correction*, p=0.0161*. **(B)** Prevalence [%] of significant depression symptoms in PLMS (n=59) and control children (n=32). Significant depression symptoms: 45.76% PLMS vs 21.88% control; Fisher’s exact test, *p=0.0401*. **(C)** Erythrocyte sedimentation rate [mm/hour] in PLMS (n=69) and control children (n=35). Median [IQR]: 8 [5–12.5] PLMS vs 7 [5–8] control; Mann-Whitney test, *p=0.016*. **(D)** Serum lipid profiles [ng/ml] in PLMS (n=70) and control children (n=35). Triglycerides (TGs) median [IQR]: 106 [76.5–145.3] PLMS vs 79 [60–100] control; Mann-Whitney test, *p=0.001*. High-density lipoproteins (HDL) mean ± SD: 48.87 ± 10.62 PLMS vs 50.63 ± 7.99 control; unpaired t-test, *p=0.3898*. Low-density lipoproteins (LDL) median [IQR]: 84 [74–105] PLMS vs 91 [82–113] control; Mann-Whitney test, *p=0.1278*. Total cholesterol (TC) median [IQR]: 150 [133.8–170.3] PLMS vs 154 [140–171] control; Mann-Whitney test, *p=0.3334*. **(E)** Pathway impact analysis of altered serum metabolites between PLMS and control children. x-axis = pathway impact values from pathway topology analysis, y-axis = –log₁₀(p) from pathway enrichment analysis. Bubble size reflects pathway impact; color intensity (from yellow to red) indicates significance. No outliers were removed. Violin plots indicate individual values supplemented with group means and IQRs. *\* p<0.05. ** p<0.01*.

### Differential miRNAs in serum of children with CT exposure

Lipid metabolism and immune system in mammals are intricately associated with circulating miRNAs. A large fraction of circulating miRNAs in mammals is carried by extracellular vesicles (EVs) released from tissues primarily involved in lipid metabolism, such as white and brown adipose tissue (Thomou et al., 2017; Diez-Roda et al., 2024). Similarly, inflammatory conditions in humans have been associated with altered levels of blood miRNAs (Nalbant & Akkaya-Ulum, 2024). Finally, HDLs, which are closely related to immunometabolic health and showed non-significant numerical decrease in PLMS children, are critical carriers of circulating miRNAs (Vickers et al., 2011). Therefore, we examined if the observed lipid alterations and indications of systemic inflammation in PLMS children are associated with changes in circulating miRNAs. Unbiased sequencing analysis of small RNAs revealed differential levels of miRNAs in the serum of PLMS children (n=48) compared to controls (n=18). There was no overall group separation based on principal component analysis (PCA) (Supplementary Fig. 2) and small RNA populations were globally comparable in PLMS and control groups (Supplementary Fig. 3). However, 49 specific miRNAs were upregulated, and 18 miRNAs were downregulated in serum of PLMS children compared to controls (with 10% FDR) (Fig. 2A). Pathway analyses of differentially expressed small RNAs showed enrichment for receptor tyrosine kinase signaling, cellular response to stress, class I MHC antigen processing and presentation, cellular response to external stimuli and Rho GTPases cycle (Fig. 2B).

**Figure 2.**
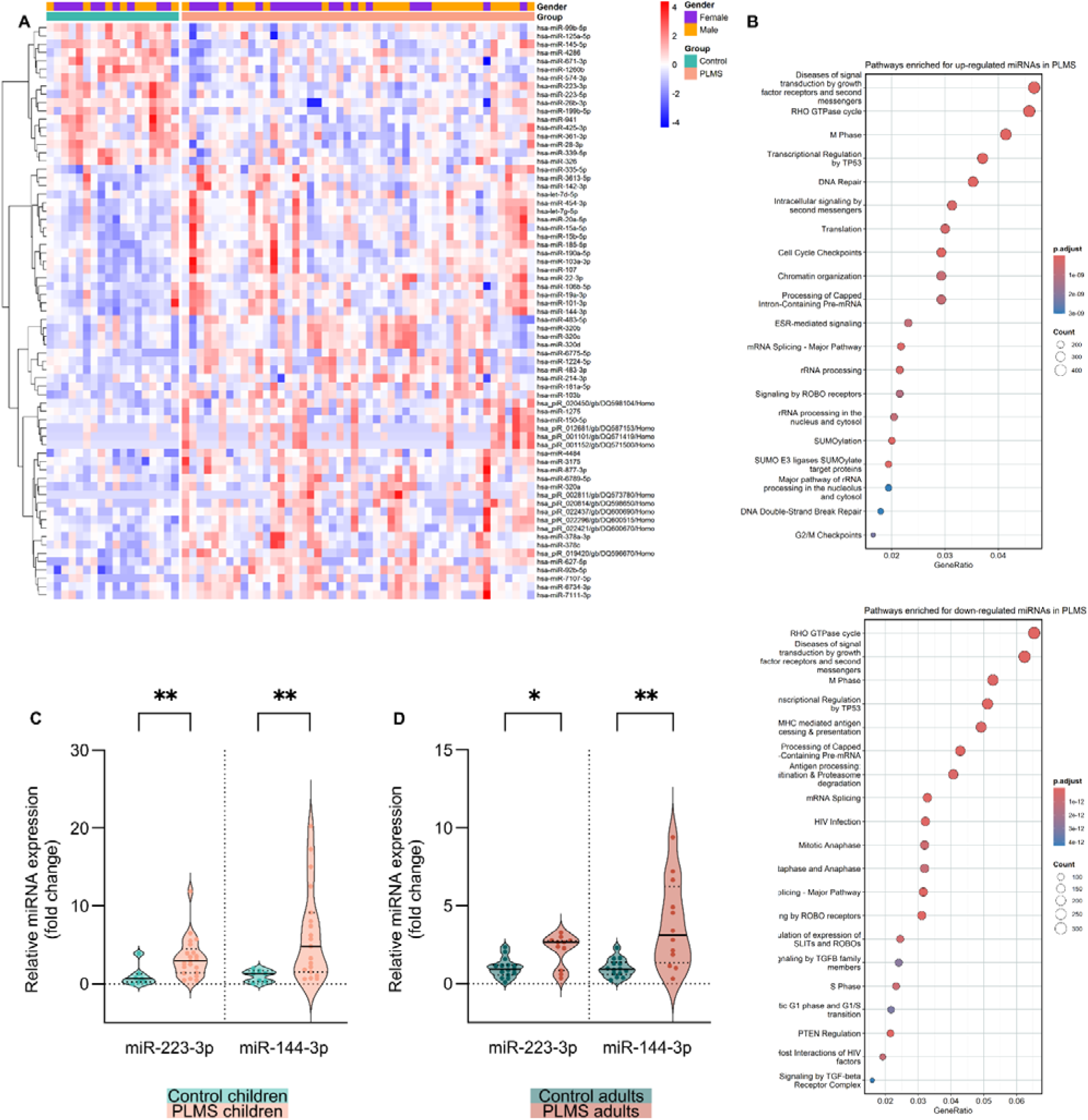
Differential level of small RNAs in the serum of PLMS children. **(A)** Heatmap of differentially expressed small RNAs in the serum of PLMS (n=48) and control (n=18) children. **(B)** Reactome pathway analysis of altered miRNAs in PLMS serum. **(C)** Serum miRNAs level in PLMS (n=22) and control (n=8) children assessed by RT-qPCR. miR-223-3p median [IQR]: 2.92 [1.39–4.5] PLMS vs 0.66 [0.21–1.27] control; Mann-Whitney test, *p=0.007*. miR-144-3p median [IQR]: 4.81 [1.51–9.15] PLMS vs 1.3 [0.3–1.63] control; Mann-Whitney test, *p=0.0045*. **(D)** Serum miRNAs level in PLMS (n=12) and control (n=15) adults assessed by RT-qPCR. miR-223-3p median [IQR]: 2.66 [0.83–2.76] PLMS vs 0.91 [0.55–1.18] control; Mann-Whitney test, *p=0.0108*. miR-144-3p mean ± SD: 3.77 ± 2.83 PLMS vs 1 ± 0.58 control; unpaired t-test with Welch’s correction, *p=0.006*. In all analyses outliers were removed using ROUT Q=1%. Violin plots indicate individual values supplemented with group means and IQRs. *\* p<0.05. ** p<0.01*.

The differential expression of miRNAs in the PLMS versus control groups was not a result of hemolysis in either of the groups. The miR-451a/miR-23a ratio, a commonly used indicator of hemolysis in serum miRNA analyses, was below the range typically considered suggestive of hemolysis in both the PLMS and control children groups (Shah et al., 2016; Supplementary Fig. 4). Similarly, sequencing-based expression of the erythrocyte-enriched miR-451a was very low overall (PLMS children: median [IQR] 17012.87 [12214.07–29450] TMM-normalized counts per million; control children: median [IQR] 17732.84 [12093.32–26679.19] TMM-normalized counts per million), indicating good sample preservation without evidence of substantial hemolysis.

We then conducted RT-qPCR assays on serum RNA to confirm the differential level of miRNAs (>1 log_2_FC in PLMS relative to control children on sequencing or <-0.5 log_2_FC). 5 candidate miRNAs were selected for follow-up, of which 2 were confirmed to be differentially regulated by targeted RT-qPCR (Fig. 2C). RT-qPCR analysis (n=22 PLMS, n=8 controls) revealed a significant increase (*p<0.01*) in serum levels of miR-223-3p (relative median [IQR] fold increase: 2.92 [1.39–4.5]) and miR-144-3p (relative median [IQR] fold increase: 4.81 [1.51–9.15]) in PLMS children compared to controls. Furthermore, there was no association between salivary cortisol and differentially expressed serum miRNAs since salivary cortisol levels were comparable (*p>0.1*) in PLMS (n=48) and control (n=20) children (Extended Data Fig. 3). Finally, given the close link between miR-223-3p and systemic lipids (Gu et al., 2022; Ye et al., 2018; You et al., 2022; Zhang et al., 2021), we additionally examined whether serum miR-223-3p levels in PLMS children varied with lipid parameters. Serum miR-223-3p in PLMS children was not significantly correlated with either of the lipid classes (HDL, LDL, TG) or total cholesterol (TC) (Extended Data Fig. 4), indicating that the observed miR-223-3p increase in PLMS children serum is not fully explained by linear associations with measured serum lipid parameters.

### Elevated miRNAs in serum of adults with CT exposure

We next examined if changes in miR-223-3p and miR-144-3p observed in serum of PLMS children can also be detected in PLMS adults. For this, we assembled a cohort of 18-25-year-old male subjects who had been exposed to PLMS when 7-12 years old and stayed in the SOS children’s village until the age of 15 (n=13). At 15, the subjects moved to SOS Youth Home, a partner organization of the SOS Children’s Village. Control subjects were age- and BMI-matched volunteer male college students (Supplementary Table 2) from the same area with no history of significant childhood or adolescence traumatic experience (n=16). Both miR-223-3p and miR-144-3p were increased in the serum of young adult males with PLMS. miR-223-3p levels were higher in adult PLMS (relative median [IQR] fold increase: 2.66 [0.83–2.76]; *p<0.05*) relative to adult controls. Similarly, miR-144-3p levels were significantly elevated in adult PLMS compared to controls (relative fold change: 3.77 ± 2.83; *p<0.01*; Fig. 2D). These results suggest that miRNA alterations associated with CT may persist until adulthood.

### Differential miRNAs in sperm of adult men with a history of complex childhood or adolescent trauma

We had previously observed miRNA changes in both serum and sperm after MSUS in mice (Gapp et al., 2014). Given the evidence from animal models that trauma-related molecular changes may extend to the germline, we next examined whether independent miRNA alterations are detectable in human sperm following CT. Due to limited samples size of the PLMS adult cohort, we assembled a separate cohort of adults with a larger sample size suitable for exploratory group-wise discovery analyses. We recruited adult men who had consulted a diagnostic facility for routine reproductive health assessments. Those who volunteered to be part of the study (n=93) were screened for CT exposure by blinded investigators via CT questionnaire (CTQ) after deposition of their seminal fluid. This questionnaire addresses six traumatic experiences (parental death, parental divorce/separation, exposure to violence or physical abuse, sexual traumatic experience, exposure to severe illness requiring hospitalization or other) before the age of 18 in adults (ECHO cohort CTQ version 01.20, November 30, 2018). Based on CTQ results, we classified the participants into three categories depending on the number of traumatic events they experienced: CTQ0 (no significant traumatic event: n=40), CTQ1 (1 significant traumatic event: n=25), CTQ2 (2 or more significant traumatic events: n=28). The perceived severity of trauma reported by the participants was comparable across different trauma types (Supplementary Figure 5). CTQ0 and CTQ2 groups were subsequently chosen for further group-wise comparisons. The two groups were comparable in terms of age and duration of sexual abstinence prior to seminal fluid collection (Supplementary Table 3). None of the participants had received professional therapy from a mental health provider. However, one participant reported being treated for schizophrenia and two participants were receiving treatment for tuberculosis. Their samples were excluded from subsequent analyses.

Small RNA sequencing revealed differential expression of several small RNAs in sperm samples from CTQ2 (n=23) compared to those from CTQ0 (n=26) men. While there was no clear group separation based on PCA (Supplementary Fig. 6) and small RNA species were comparable (Supplementary Fig. 7), 46 small RNAs were upregulated, and 1 small RNA was downregulated in sperm of CTQ2 compared to CTQ0 men (with 10% FDR) (Fig. 3A). Pathway analysis of differentially expressed miRNAs showed enrichment for pathways relevant to receptor tyrosine kinases, Rho GTPases signaling, and transcriptional regulation by TP53 (Fig. 3B). We selected miRNAs increased in CTQ2 sperm by a factor of >1 log2FC compared to CTQ0 for confirmation by RT-qPCR assays (n=22 CTQ2 vs n=26 CTQ0). 5 candidate miRNAs were selected for follow-up, of which 2 were confirmed by targeted RT-qPCR (Fig. 3C). In individuals with CTQ2, miR-223-3p and miR-145-5p were significantly increased compared to individuals with no trauma (CTQ0). miR-223-3p levels were higher in CTQ2 (relative median [IQR] fold increase: 1.52 [0.48–6.81] vs CTQ0, *p<0.05*). Similarly, miR-145-5p levels were significantly elevated in CTQ2 (relative median [IQR] fold increase: 2.54 [0.99–4.83] vs CTQ0; *p<0.01*) compared to CTQ0.

**Figure 3.**
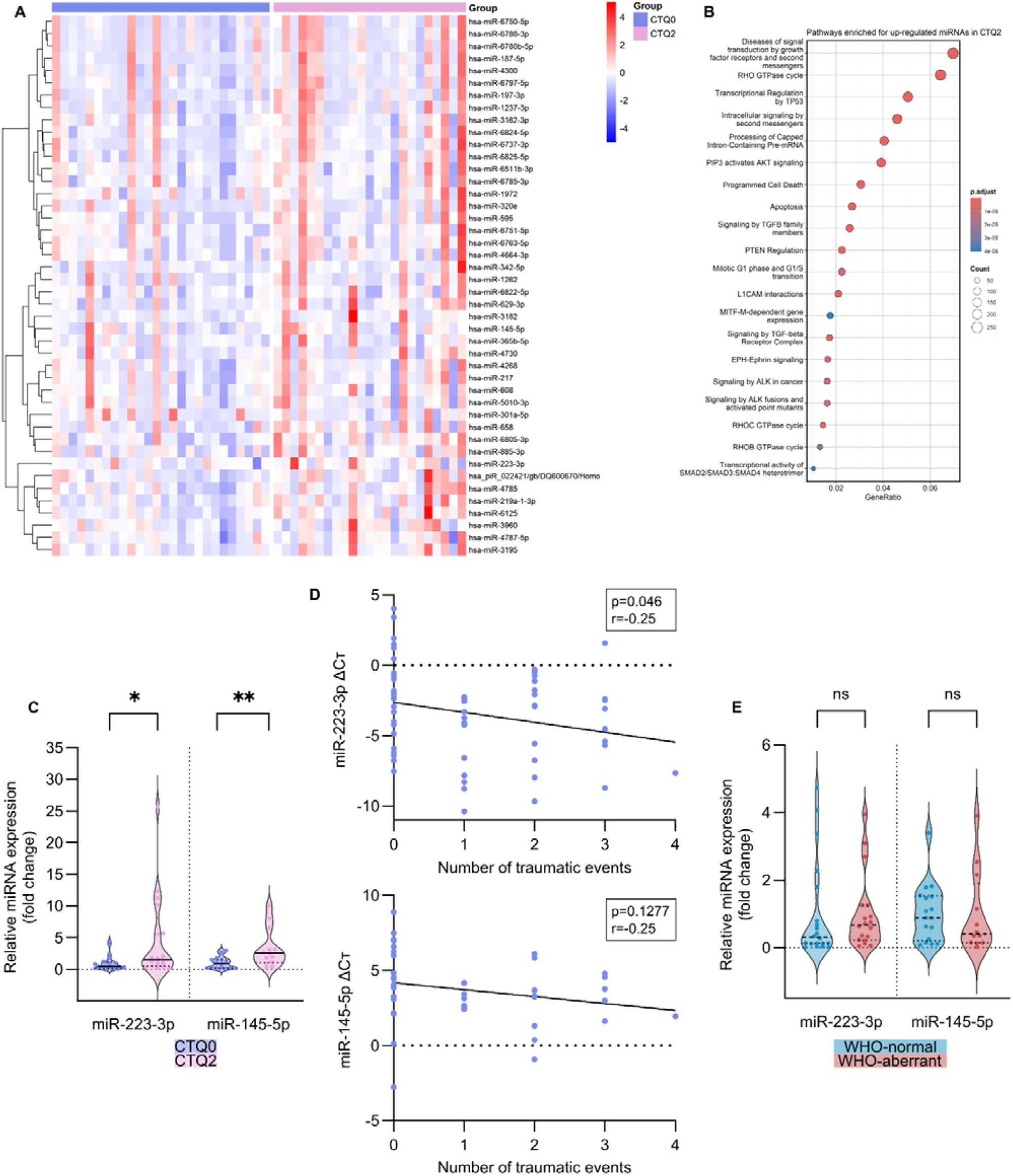
Differential level of sperm small RNAs in adult men with complex CT. **(A)** Heatmap of differentially expressed small RNAs in the sperm of adult men with two or more CT events (CTQ2, n=23) and those with no CT (CTQ0, n=26). **(B)** Reactome pathway analysis of the altered miRNAs in CTQ2 sperm. **(C)** Sperm miRNAs level in CTQ2 (n=22) and CTQ0 (n=26) men assessed by RT-qPCR. miR-223-3p median [IQR]: 1.52 [0.48–6.81] CTQ2 vs 0.44 [0.12–1.19] CTQ0; Mann-Whitney test, *p=0.0188*. miR-145-5p median [IQR]: 2.54 [0.99–4.83] CTQ2 vs 0.89 [0.2–1.51] CTQ0; Mann-Whitney test, *p=0.0044*. **(D)** Correlation of sperm miRNAs level assessed by RT-qPCRs with the number of traumatic events (n=62). miR-223-3p: Pearson correlation, *p=0.046, r=-0.25*. miR-145-5p: Pearson correlation, *p=0.1277, r=-0.25.* **(E)** Sperm miRNAs level in WHO-normal (n=25) and WHO-aberrant (n=23) samples. miR-223-3p median [IQR]: 0.66 [0.22–1.25] WHO-aberrant vs 0.3 [(0.12–1.55] WHO-normal; Mann-Whitney test, *p=0.2765*. miR-145-5p median [IQR]: 0.4 [0.14–1.9] WHO-aberrant vs 0.87 [0.2–1.52] WHO-normal; Mann-Whitney test, *p=0.6788*. In all analyses outliers were removed using ROUT Q=1%. Violin plots indicate individual values supplemented with group means and IQRs. *\* p<0.05. ** p<0.01*.

We additionally examined correlations between the number of traumatic exposures and delta cycle threshold (dCt) values for sperm miR-223-3p and miR-145-5p. A significant inverse correlation was observed for miR-223-3p (*p<0.05*, r=-0.25), indicating that a greater number of traumatic exposures was associated with higher sperm miR-223-3p expression (Fig. 3D). To further support the differential miRNA discovery pipeline - small RNA sequencing followed by RT-qPCR validation - we additionally examined the miRNAs that showed the most stable expression across CTQ2 and CTQ0 sperm samples in the sequencing dataset via RT-qPCR (Extended Data Fig. 5). RT-qPCR analyses confirmed that these miRNAs were comparably expressed between the CTQ0 and CTQ2 groups.

We then tested miR-34c and miR-449a-5p, two miRNAs previously reported as sperm signatures of childhood adversity (Dickson et al., 2018) and observed a trend (*p<0.1*) towards a decrease in CTQ2 sperm (Extended Data Fig. 6). However, sequencing-based sperm level of miR-34c and miR-449a-5p did not show a significant correlation with CTQ scores (Supplementary Fig. 8). Finally, to extend comparison with recent sperm miRNA studies of childhood adversity, we quantified by RT-qPCR five miRNAs previously reported to be associated with childhood maltreatment by Tuulari et al. (2025). Among these, miR-141-3p was significantly downregulated in CTQ2 sperm relative to CTQ0 sperm (*p<0.05*), whereas miR-21-5p, miR-29c-3p, miR-148a-3p, and miR-101-3p showed comparable expression between these groups (Extended Data Fig. 7).

We also assessed sperm quality in collected seminal fluid samples to confirm that the observed association between miRNA changes and CTQ2 was not confounded. We categorized samples into ‘normal’ or ‘aberrant’ using WHO criteria that classify seminal fluid based on sperm count, concentration, morphology, shape, pH, and liquefaction time. The proportion of normal versus aberrant samples was comparable between CTQ0 and CTQ2 samples (Supplementary Fig. 9). Further, the level of miR-223-3p and miR-145-5p was comparable in individuals with normal and aberrant sperm (Fig. 3E). These results suggest that the increased level of miR-223-3p and miR-145-5p in sperm of CTQ2 men is not confounded by low seminal fluid quality.

### Potential origins of differentially expressed sperm miRNAs

While the exact mechanisms of how adult human sperm acquires miRNAs remain elusive, the miRNA content of adult mouse sperm has been shown to largely depend on uptake of miRNAs from the blood and epididymal EVs and HDLs (Nixon et al., 2015). During sperm maturation, the sperm genome undergoes critical modification, in particular, chromatin condensation, resulting in transcriptional silencing (Rathke et al., 2014). Consequently, a substantial fraction of sperm miRNAs is acquired exogenously, initially from blood EVs and HDLs during early stages of sperm biogenesis, and subsequently via epididymal EVs and HDLs during epididymal transit (Sullivan, 2015; James et al., 2020).

We next checked if the increased miR-223-3p and miR-145-5p in sperm of CTQ2 men are related to seminal fluid, which contains abundant epididymal EVs (Saez et al., 2013). For that, we quantified and compared the level of miR-223-3p and miR-145-5p in seminal fluid of CTQ2 (n=9) and CTQ0 (n=14) men. miR-145-5p had a trend (*p<0.1*) towards an increase (relative median [IQR] fold increase: 2.83 [0.88–4.36] CTQ2 vs CTQ0) in CTQ2 men compared to CTQ0, whereas miR-223-3p was comparable (*p>0.1*) between the two groups (Fig. 4A).

**Figure 4.**
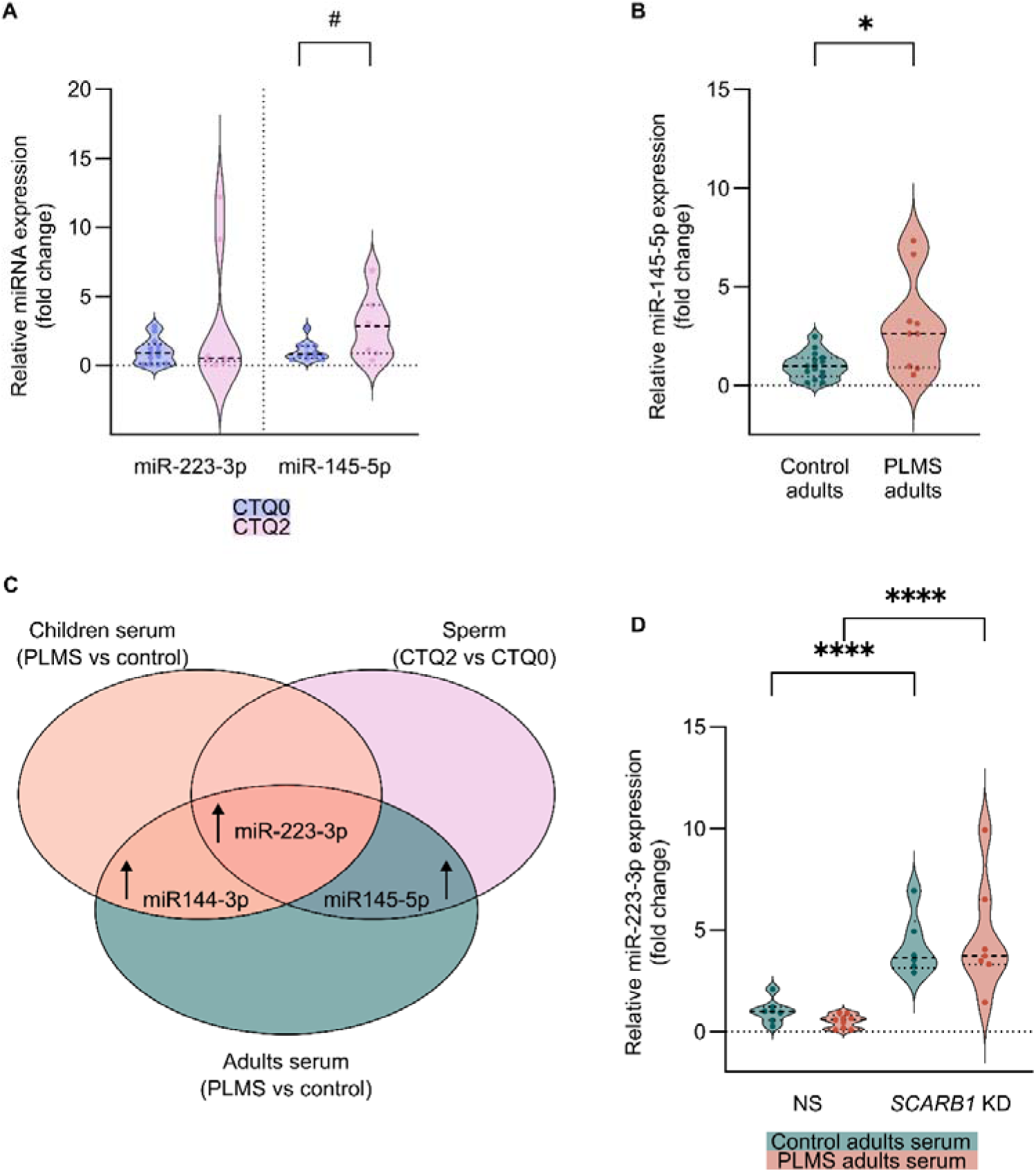
Potential origins of differential sperm miRNAs after CT. **(A)** miRNAs level in the seminal fluid of CTQ2 (n=14) and CTQ0 men (n=9) assessed by RT-qPCR. miR-223-3p median [IQR]: 0.53 [0.27–6.99] CTQ2 vs 0.89 [0.09–1.54] CTQ0; Mann-Whitney test, *p=0.9155*. miR-145-5p median [IQR]: 2.83 [0.88–4.36] CTQ2 vs 0.79 [0.52–1.42] CTQ0; Mann-Whitney test, *p=0.0831*. **(B)** Serum miR-145-5p level in PLMS (n=15) and control (n=9) adults assessed by RT-qPCR. Mean ± SD: 3.1 ± 2.43 PLMS vs 1.0 ± 0.67 control; unpaired t-test with Welch’s correction, *p=0.0325*. **(C)** Overlap between differentially expressed miRNAs in PLMS children serum, PLMS adult serum, and CTQ2 adult sperm. **(D)** miR-223-3p expression in GC-1 cells transfected with non-silencing control siRNA (NS) or siRNA targeting SR-B1 coding gene (*SCARB1* KD) upon treatment with serum from adult PLMS or control subjects (NS PLMS serum n=9; NS control serum n=7; *SCARB1* KD PLMS serum n=8; *SCRAB1* KD control serum n=6). Mean ± SD: 1.00 ± 0.57 NS (control serum), 0.50 ± 0.33 NS (PLMS serum), 4.20 ± 1.51 *SCARB1* KD (control serum), 4.62 ± 2.77 (PLMS serum); two-way ANOVA with Tukey’s post-hoc test, *p=0.949* (PLMS vs control), *p<0.0001* (*SCARB1* KD vs NS), *p=0.432* (interaction). In all analyses outliers were removed using ROUT Q=1%. Violin plots indicate individual values supplemented with group means and IQRs. *# p<0.1. * p<0.05. **** p<0.0001*.

We then examined miRNAs elevated in CTQ2 sperm in the serum of PLMS adults. In addition to the aforementioned increase in miR-223-3p (Fig. 2D), miR-145-5p was also increased in adult PLMS serum (relative fold increase: 3.1 ± 2.43 vs adult controls; Fig. 4B). Moreover, miR-320e, found elevated in the CTQ2 sperm in the sequencing data, was also increased in the serum of PLMS adults (relative median [IQR] fold increase: 5.08 [0.67–8.57] adult PLMS vs adult control; Supplementary Fig. 10).

Notably, miR-223-3p was the only miRNA upregulated in sera of PLMS children and adults, as well as in the sperm of CTQ2 adult men (Fig. 4C). miR-223-3p is also unique because it is carried by HDL in circulation and targets scavenger receptor class B type 1 (SR-B1), the receptor for HDL (Shen et al., 2018; van Niel et al., 2018). We examined if miR-223-3p increase in sperm of CTQ2 men could be acquired at an earlier stage of spermiogenesis, when spermatogonial stem cells are in direct contact with blood EVs and HDLs. To test this hypothesis, we treated GC-1 type B spermatogonia-like cells (GC-1) with serum from PLMS adults or controls. Treatment of GC-1 cells with adult PLMS serum did not alter the level of miR-223-3p compared to treatment with control serum. However, when SR-B1 was knocked down (Supplementary Fig. 11), miR-223-3p expression was significantly increased in GC-1 cells treated with either control (relative fold increase after SR-B1 knockdown: 4.20±1.51) or adult PLMS serum (relative fold increase after SR-B1 knockdown: 4.62±2.77) (Fig. 4D). On the contrary, the SR-B1 knockdown did not alter the expression of miR-145-5p in either the adult control or adult PLMS serum treatment groups (Extended Data Fig. 8). These results suggest that the miRNA profile of spermatogonia-like cells may be altered in a miRNA-specific manner through mechanisms involving HDL receptor signaling, independent of whether the cells were exposed to adult PLMS or control serum.

### Alteration of miR-223-3p in a mouse model of early postnatal traumatic stress associated with transgenerational phenotypes

A key question arising from the observation that miRNAs are altered in the sperm of adult men with complex trauma (CTQ2) compared to controls (CTQ0) is whether these changes may contribute to the intergenerational transmission of trauma-associated phenotypes. The lack of assessments in the next generation, however, prevents any causal or correlative inference between sperm miRNA and intergenerational effects after CT. Therefore, we examined miR-223-3p and miR-145-5p in the MSUS mouse model, which exhibits transgenerational transmission of behavioral and metabolic phenotypes via the patriline (Franklin et al., 2010; Gapp et al., 2014; Gapp et al., 2017; van Steenwyk et al., 2018; Boscardin et al., 2022) (Fig. 5A). miR-223-3p and miR-145-5p levels were lower in sperm of 3-month-old MSUS males (n=12) compared to age-matched control males (n=7) (relative fold decrease miR-223-3p: 0.18±0.06 MSUS vs control, *p<0.01*; relative median [IQR] fold decrease miR-145-5p: 0.23 [0.2–0.55] MSUS vs control, *p<0.01*) (Fig. 5B). Although not directly comparable to human samples, the differential regulation of miR-223-3p and miR-145-5p in the MSUS mouse model suggests a potential relevance of these miRNAs to intergenerational effects after paternal CT. To broaden the cross-species comparison of sperm miRNA signatures of early life trauma, we also examined miR-16-5p and miR-375-5p, previously found to be elevated in MSUS sperm (Gapp et al., 2014; Jawaid et al., 2020), in human samples. Both miR-16-5p and miR-375-5p were significantly lower in sperm from CTQ2 men (n=23) compared to CTQ0 (n=35) men (relative median [IQR] fold decrease miR-16-5p: 0.53 [0.32–1.15] CTQ2 vs CTQ, *p<0.05*; relative median [IQR] fold decrease miR-375-5p: 0.38 [0.25–0.68] CTQ2 vs CTQ0, *p<0.01)* (Fig. 5C).

**Figure 5.**
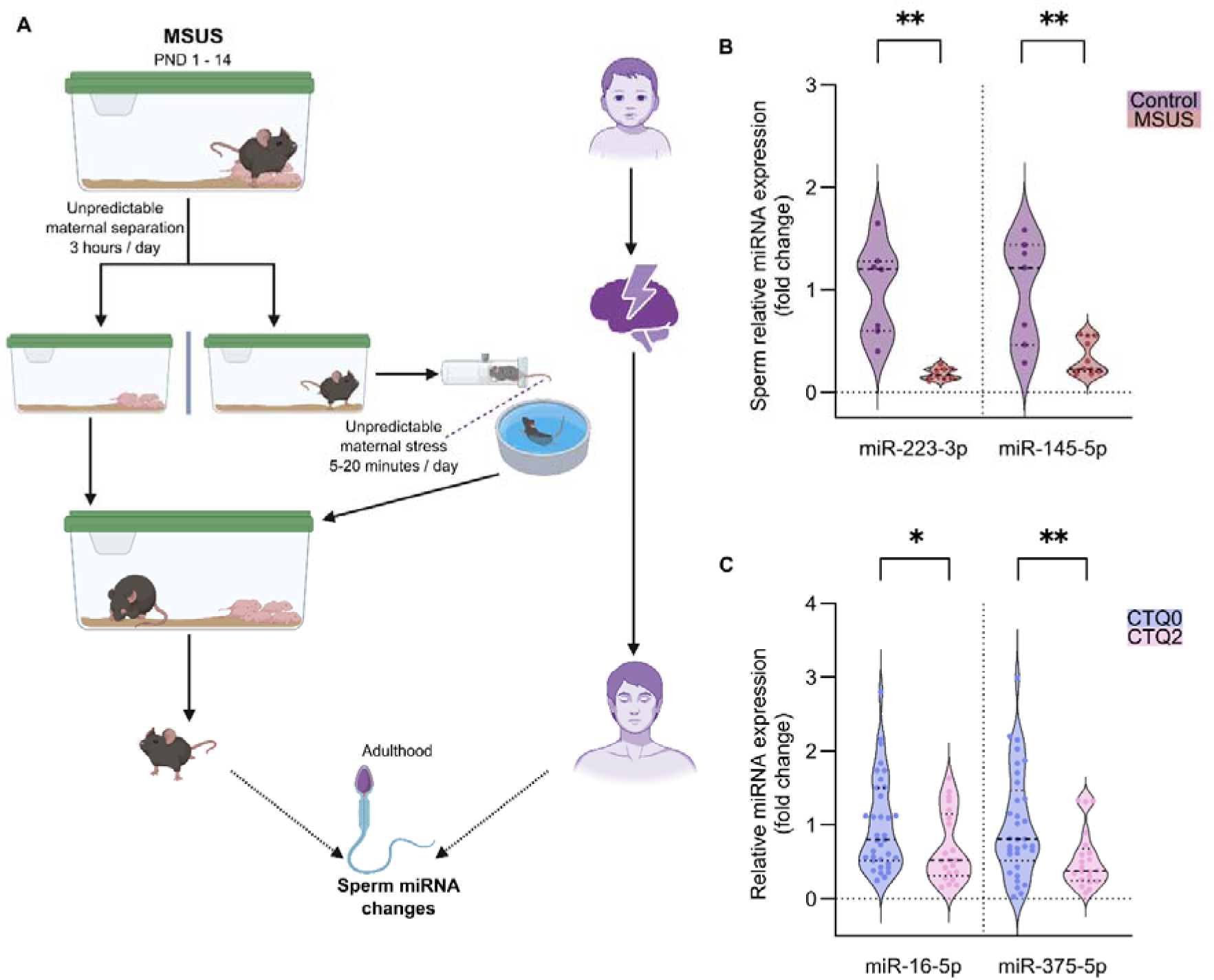
Cross-species assessment of sperm miRNAs after CT in mice and humans **(A)** Graphical illustration of the MSUS model in mice (left) and CT in humans (right). **(B)** miRNAs levels in the sperm of MSUS (n=12) and control (n=7) males assessed by RT-qPCR. miR-223-3p mean ± SD: 0.18 ± 0.06 MSUS vs 1 ± 0.45 control; unpaired t-test with Welch’s correction, *p=0.003*. miR-145-5p median [IQR]: 0.23 [0.2–0.55] MSUS vs 1.21 [0.46–1.44] control; Mann-Whitney test, *p=0.0059*. **(C)** Effect of complex trauma (CTQ2) on the level of miRNAs in human sperm previously found to be altered in MSUS sperm. Sperm miRNAs level in CTQ2 men (n=23) and CTQ0 men (n=35) assessed by RT-qPCR. miR-16-5p median [IQR]: 0.53 [0.32–1.15] CTQ2 vs 0.8 [0.52–1.51] CTQ0; Mann-Whitney test, *p=0.0411*. miR-375-5p median [IQR]: 0.38 [0.25–0.68] CTQ2 vs 0.81 [0.52–1.47] CTQ0; Mann-Whitney test, *p=0.0077*. In all analyses outliers were removed using ROUT Q=1%. Violin plots indicate individual values supplemented with group means and IQRs. *\* p<0.05. ** p<0.01*.

We further tested if decreased miR-223-3p and miR-145-5p in MSUS sperm would be consistent with a similar pattern of expression in serum. However, miR-223-3p, as well as miR-145-5p levels, were comparable in MSUS versus control serum (Extended Data Fig. 9). Furthermore, no significant correlations were found between serum and sperm miR-145-5p and only a trend (*p<0.1*) was noted for miR-223-3p (Supplementary Fig. 12).

### Mimicking miR-223-3p increase in 1-cell mouse embryos alters brain and adipose miRNA expression and metabolism in the resulting mice

We next examined if manipulating miR-223-3p level in the early embryo will have molecular effects in the resulting animals later in life. We delivered miR-223-3p mimic into fertilized control oocytes and derived adult animals from the embryos (Fig. 6A). The embryos were obtained from the mating of superovulated naïve C57BL/6J females with naïve C57BL/6J males. Following miRNA mimic delivery, the embryos were implanted into foster CD1 mothers, and the resulting mice were monitored till adulthood and sacrificed when 1 year-old for tissue harvest. The concentration of the miRNA mimic was determined based on pilot experiments in GC-1 cells, where the miR-223-3p mimic effectively suppressed its targets (Supplementary Fig. 13). The analyses in the adult mice resulting from the embryonic miRNA mimic delivery was limited to females as a number sufficient for statistical comparisons for male embryos was not reached. miR-223-3p was significantly increased in the brain of the resulting female mice when adult compared to females from control (relative fold increase: 1.22±0.14 miR-223-3p mimic vs control; *p<0.05*; Fig. 6B). Furthermore, miR-223-3p quantification in the adipose tissue revealed an increase in only white adipose tissue of the resulting females from miR-223-3p mimic (relative fold increase: 1.9±0.1 miR-223-3p mimic vs control; *p<0.0001*; Fig. 6B).

**Figure 6.**
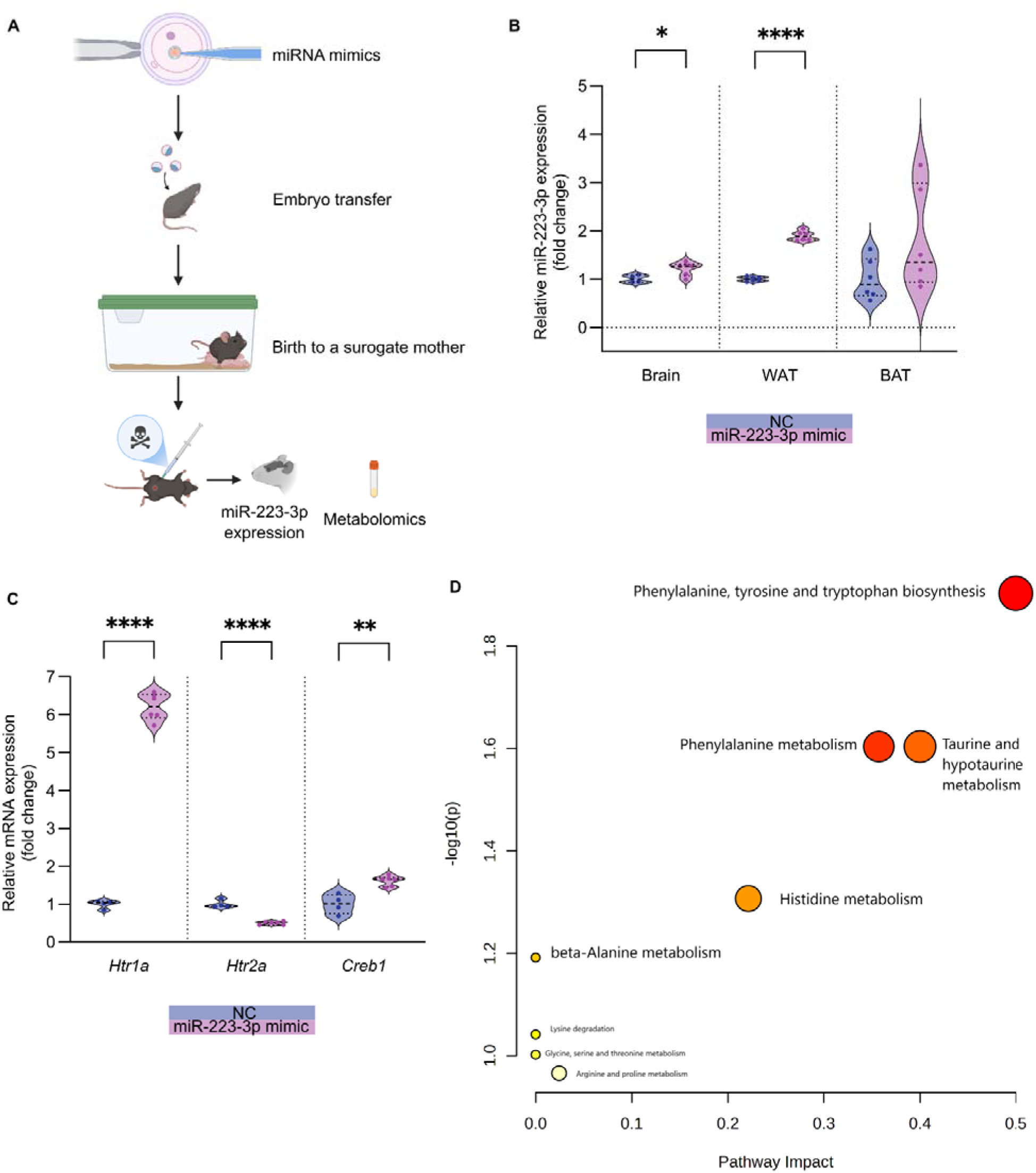
Molecular effects of embryonic miR-223-3p mimic injection on the resulting animals’ brain, adipose tissue, and serum miR-223-3p expression and serum metabolome. **(A)** Scheme of miR-223-3p mimic delivery into 1-cell mouse embryos. **(B)** miR-223-3p level in the brain, white adipose tissue (WAT), and brown adipose tissue (BAT) of mice developed from embryos treated with negative control (NC n=6) or miR-223-3p mimic (n=6) assessed by RT-qPCR. Brain mean ± SD: 1.22 ± 0.14 mimic vs 1 ± 0.08 NC; unpaired t-test, *p=0.0269*. WAT mean ± SD: 1.9 ± 0.1 mimic vs 1 ± 0.05 NC; unpaired t-test, *p<0.0001*. BAT mean ± SD: 1.8 ± 1.1 mimic vs 1 ± 0.42 NC; unpaired t-test, *p=0.1220*. **(C)** Expression of serotonin receptors and plasticity-related genes in the brain of mice developed from embryos treated with NC (n=6) or miR-223-3p mimic (n=6) assessed by RT-qPCR. *Htr1a* mean ± SD: 6.21 ± 0.35 mimic vs 1 ± 0.11 NC; unpaired t-test, *p<0.0001*. *Htr2a* mean ± SD: 0.5 ± 0.04 mimic vs 1 ± 0.1 NC; unpaired t-test, *p<0.0001*. *Creb1* mean ± SD: 1.61 ± 0.13 mimic vs 1 ± 0.26 NC; unpaired t-test, *p=0.0011*. **(D)** Pathway impact analysis of altered serum metabolites between mice generated from miR-223-3p mimic and NC embryos. x-axis = pathway impact values from pathway topology analysis, y-axis = –log₁₀(p) from pathway enrichment analysis. Bubble size reflects pathway impact; color intensity (from yellow to red) indicates significance. In all analyses outliers were removed using ROUT Q=1%. Violin plots indicate individual values supplemented with group means and IQRs. *\* p<0.05. ** p<0.01. **** p<0.0001*.

To assess the potential functional consequences of early embryonic increase in miR-223-3p, we also compared expression of key genes involved in major plasticity cascades and serotonergic signaling in the brain, as well as circulating metabolome of the 1-year-old resulting mice (Kraus et al., 2017; Nestler et al., 2002). Expression of *Htr1a and Htr2a*, genes encoding serotonin receptors; and *Creb1,* encoding plasticity-associated transcription factor, was altered in brain of adult mice resulting from the embryonic delivery of miR-223-3p mimic (relative fold increase *Htr1a*: 6.21±0.35 miR-223-3p mimic vs control; *p<0.0001*; *Creb1*: 1.61±0.13 miR-223-3p mimic vs control; *p<0.001, relative fold decrease Htr2a*: 0.5±0.04 miR-223-3p mimic vs control; *p<0.0001)* (Fig. 6C). Furthermore, unbiased metabolomics analyses of serum from the resulting animals showed enrichment for metabolites associated with phenylalanine, tyrosine and tryptophan biosynthesis, phenylalanine metabolism and taurine and hypotaurine metabolism, suggesting altered metabolome (Fig. 6D).

## DISCUSSION

This study identifies miRNA alterations associated with CT across multiple biological compartments, including serum and sperm, with limited but notable overlap. Distinct miRNA alterations were present in the serum of children shortly after CT exposure via PLMS, with similar changes persisting into adulthood. Differential miRNA expression was also observed in sperm of men with complex CT. Notably, miR-223-3p, a key regulator of cholesterol biosynthesis, emerged as a convergent signal across cohorts and tissues, suggesting it may reflect a shared molecular response to CT. Similarly, miRNAs altered in sperm of men with complex trauma were dysregulated in MSUS mice, a model of postnatal traumatic stress with known transgenerational effects. As proof of concept, we delivered a miR-223-3p mimic into one-cell embryos from naïve mice. The resulting adult female mice showed upregulation of miR-223-3p in brain and white adipose tissue accompanied with changes in serum metabolome and genes related to serotonergic signaling. These findings broadly support our hypotheses that CT is associated with specific miRNA signatures in serum and sperm, and that CT-related miRNAs may be relevant to pathways implicated in intergenerational effects.

This study is among the first systematic and unbiased assessments of miRNA signatures associated with CT in humans in both serum and sperm. Serum miRNAs were analyzed in children and young adults to evaluate the persistence of CT-induced alterations. In parallel, sperm miRNA profiles were examined in an independent cohort of adult men with complex trauma. Importantly, all human cohorts originated from a population with high consanguinity, thereby reducing genetic heterogeneity within the cohort. While this does not recapitulate the uniformity of inbred mouse models, it may facilitate cross-species comparisons. Overlapping miRNAs were altered, albeit in contrasting direction, in the sperm of mice exposed to an ethologically relevant model of CT based on MSUS, which could reflect inter-species differences.

Previous studies examining sperm miRNA signatures associated with childhood adversity in humans have reported altered expression of miR-34c-5p (Dickson et al., 2018; Tuulari et al., 2025) and miR-449a-5p (Dickson et al., 2018). In our study, unbiased small RNA sequencing did not identify differential level of miR-34c-5p or miR-449a-5p in sperm of CTQ2 men. Furthermore, unlike previously reported associations, we did not see a correlation between the RNAseq-based sperm expression of miR-34c-5p and miR-449-5p and CTQ scores. However, targeted RT-qPCR assays showed a trend (*p*<0.1) towards lower miR-34c-5p and miR-449-5p level in sperm of CTQ2 men compared to CTQ0. A possible explanation for this discrepancy is that earlier studies on sperm small RNA signatures of CT used slightly smaller cohorts (n=30 in Tuulari et al. and n=28 in Dickson et al.) for correlation analysis compared to our cohort (n=66). Additionally, these studies were conducted in Western populations, and differences related to ethnicity and lifestyle may have influenced baseline sperm levels of these miRNAs. Methodological differences may as well partly account for different results. Dickson et al. used microarray technology for transcriptomic analysis, which has limited sensitivity for detecting differentially expressed non-coding RNAs compared to small RNA sequencing (Dickson et al., 2018). In contrast, while the Tuulari study employed RNA sequencing for initial profiling, it did not report RT-qPCR confirmations (Tuulari et al., 2025).

We employed an unbiased approach for miRNA profiling in sperm from men with varying CT level and confirmed miRNA changes by RT-qPCR in the same RNA samples, providing confidence in the results. This approach combines an unbiased and high-throughput discovery platform that is corrected for FDR followed by RT-qPCRs that offer targeted confirmations. This approach also revealed a high but not complete degree of concordance between sequencing and qPCR results. The direction and magnitude of change were consistent across both approaches for sperm but not for serum miR-223-3p. This partial concordance between sequencing and qPCR validation underscores the importance of integrating both approaches in future studies combining serum and sperm ncRNA signatures of CT and related exposures.

Furthermore, we extended our analyses to serum miRNAs in both children and adults from the PLMS cohort, providing independent support for our methodological framework. Prior studies relying solely on retrospective self-report measures of CT are susceptible to recall bias due to the temporal gap between exposure and sperm collection. In contrast, the identification of consistent miRNA alterations in serum across cohorts with both recent and remote CT exposure, together with partially overlapping signatures observed in sperm from an independent cohort, supports a biological association between CT and miRNA changes across tissues.

To further strengthen this framework, we implemented an alternative discovery and validation pipeline, whereby miRNAs identified as differentially expressed in CTQ2 versus CTQ0 sperm by sequencing were subsequently tested in serum from PLMS adults. This approach expanded the set of validated trauma-associated miRNAs in adult PLMS serum to include miR-145-5p and miR-320e. Taken together, these findings support the robustness of CT-associated miRNA signatures, while indicating partial overlap across tissues and reinforcing their potential relevance as biomarkers of early-life adversity. Importantly, the serum and sperm analyses were conducted in independent cohorts and should be interpreted as reflecting parallel but not directly linked biological processes. While overlap of specific miRNAs may suggest potential shared regulation, the current study does not establish direct transfer or correspondence between circulating and germline miRNAs in humans. Future studies with matched sampling will be required to address this question.

The miRNAs differentially regulated by CT in our study are closely linked to lipid metabolism. miR-223-3p, consistently altered in serum following PLMS and in sperm after complex trauma, plays a central role in cholesterol biogenesis (Vickers et al., 2014). miR-223 alterations are associated with atherosclerosis, insulin resistance, non-alcoholic fatty liver disease and non-alcoholic steatohepatitis (Gu et al., 2022; Ye et al., 2018; You et al., 2022; Zhang et al., 2021). It regulates several key proteins involved in lipid metabolism, including fatty acid synthase and SR-B1 (Vickers et al., 2014). Through these and possibly other regulatory mechanisms, elevated levels of miR-223 may influence lipid-associated pathways, such as cholesterol content in sperm. Importantly, miR-223 is one of the most abundant miRNAs associated with HDL in circulation and regulates the post-transcriptional expression of the HDL receptor, SR-B1 (Tabet et al., 2014; Cuesta Torres et al., 2019). This suggests that extracellular and intracellular miR-223, especially in tissues like testis, may be interconnected via self-regulatory loops sensitive to lipid signaling. This interplay is supported by our *in vitro* “serum on cells” experiment, in which miR-223-3p in GC-1 spermatogonia-like cells significantly increased following SR-B1 knockdown. Notably, SR-B1 expression is known to be reduced in sperm of men with obesity (Calderón et al., 2019), potentially affecting miR-223-3p but also other HDL-associated miRNAs. To verify that, we tested the expression of miR-145-5p, elevated in sperm of CTQ2 men, and previously shown to be altered in the sperm of HFD-exposed mice (Fullston et al., 2016). However, SR-B1 knockdown did not impact the GC-1 levels of miR-145-5p indicating the potential link between CT, metabolic state, and sperm miRNAs may be selective and not global.

Changes in sperm ncRNAs have previously been linked to intergenerational phenotypes of early-life stress in mice (Gapp et al., 2014). We provide proof-of-concept evidence that miR-223-3p, altered in human sperm after CT, can influence molecular phenotypes later in life when elevated at the zygote stage in mice. miR-223-3p is undetectable in oocytes and is therefore likely delivered to the embryo via sperm (Battaglia et al., 2016). We tested for the effect of early embryonic increase in miR-223-3p by delivering miR-223-3p mimic into 1-cell embryos. Notably, in the resulting animals, miR-223-3p expression was significantly increased in the brain and white adipose tissue with accompanying changes in brain expression of genes encoding serotonergic receptors and alterations in the serum metabolome. These findings were paralleled by consistent changes in lipid-associated miRNAs across cohorts, including the serum of PLMS children and adults, as well as the sperm samples from the cohort of adult men. This convergence suggests a shared lipid signaling axis that may be relevant to CT-associated molecular responses and could contribute to mechanisms implicated in intergenerational effects.

However, several aspects of our findings need to be carefully interpreted when considering their broader implications. A key limitation of our study is the absence of miRNA assessments in both serum and sperm samples from the same individuals. Although analyzing sperm miRNAs in PLMS adults was a possibility, the relatively small sample size limited statistical power and prevented these analyses. Conversely, blood collection was not permitted from the CTQ cohort, as participants were primarily focused on fertility assessments and were generally in good health. Despite these constraints, the partially overlapping miRNA alterations observed in serum of PLMS children, PLMS adults and CTQ2 men, suggest a potential relevance of these miRNAs in CT-induced intergenerational phenotypes. Comparative interspecies analyses in a mouse model with marked transgenerational phenotypes, along with observed changes in animals resulting from embryonic delivery of miR-223-3p mimic, support this hypothesis.

Another limitation is the relatively small sample size across cohorts. In particular, the adult PLMS cohort (n=16 PLMS adults, n=13 controls) did not have sample size large enough for identifying differential signatures. However, both discovery cohorts, the PLMS children cohort (n=70 PLMS children, n=35 control children) and adult men CTQ cohort (n=93) are, to our knowledge, among the largest reported in this research area. In specific group-wise comparisons for miRNAs discovery, serum samples from n=48 PLMS children were analyzed against n=18 controls, whereas sperm samples from n=23 CTQ2 men were analyzed against n=26 controls. Due to the small sample size for the PLMS adult cohort, it was only used for targeted miRNA validations via RT-qPCR. The PLMS children serum and CTQ sperm cohorts served as the primary discovery datasets, whereas the smaller adult PLMS cohort was used for targeted validation rather than unbiased discovery. Accordingly, human discovery analyses should be interpreted as exploratory and hypothesis-generating until replicated in larger matched cohorts. This could also be the reason for some unexpected large effect sizes, especially in the adult PLMS cohort, which should be interpreted with caution.

Further, sample sizes vary widely across analyses in our study (Extended Data Figure 1). This is particularly evident in the comparison of serum from PLMS and control children, for which there were 66 samples (n=48 PLMS children, n=18 control children) for small RNA sequencing and only 30 samples (n=22 PLMS children, n=8 control children) for RT-qPCR assays. This difference is primarily due to the limited availability and low integrity of RNA from serum samples and logistical challenges that necessitated RT-qPCR analyses at two different time points. RNA extraction from PLMS serum was conducted in early 2020, but due to COVID-19 pandemic, small RNA sequencing could not be carried out until late 2021. In the interim, we employed a candidate approach to quantify miRNA changes by RT-qPCRs (Jawaid et al., 2020). This resulted in significant sample loss, further reducing the availability of material for sequencing and subsequent RT-qPCR assays.

Finally, the lack of longitudinal data is a limitation. For the very reason, BMI was used as a proxy indicator of metabolic health as long-term assessments of nutritional status, physical activity, and other more robust metabolic parameters were not available. The utility of BMI as a sole indicator of nutritional status and metabolic health is questionable and should be considered as a limitation (Byker Shanks et al. 2025). However, we continue to monitor the health and developmental trajectories of PLMS children, who are now between 12 and 17 years old with the long-term objective to continue the assessments in adulthood, including sperm analyses in male participants.

In conclusion, through comprehensive, unbiased analyses and validation across three independent cohorts characterized by high genetic consanguinity, this study has identified distinct serum and sperm miRNA signatures associated with CT. Notably, sperm miRNA alterations show interspecies relatability. Experimental manipulation of miR-223-3p, consistently altered in both human serum and sperm following CT, in mouse zygotes resulted in specific metabolomic and transcriptomic alterations. Collectively, these findings identify serum and sperm miRNAs as molecular correlates of CT with potential implications for intergenerational biology and nominate miR-223-3p as a convergent biomarker candidate.

## Supporting information

Supplementary statistics

## Data Availability

All raw data is available upon request

## Acknowledgments

This work was supported by funds and grants awarded to IMM by the Swiss National Science Foundation (NF 31003A_135715 and NF 31003A_175742/1), ETH grants (ETH-10 15-2 and ETH-17 13-2), the National Centre of Competence in Research (NCCR) RNA&Disease funded by the Swiss National Science Foundation (grant number 182880/Phase 2 and 205601/Phase 3), the Hochschulmedizin Flagship Project “STRESS”, the European Union Horizon 2020 Research Innovation Program EarlyCause (Grant number 848158), the European Union projects FAMILY and HappyMums funded by the Swiss State Secretariat for Education, Research and Innovation (SERI), the FreeNovation grant from Novartis Forschungsstiftung and the Escher Family Fund, and grants to AJ from the Polish National Science Center (SONATA; DEC-2020/39/D/NZ3/01887) and ERA Net NEURON (NeuronC2/III/39/2024/MUSEACE). AJ, MG, and IG were employed for a part of the project by BRAINCITY: Center of Excellence for Neural Plasticity and Brain Disorders. BRAINCITY project was carried out within the International Research Agenda Program (IRAP) co-financed by the European Union under European Regional Development Funds.

We are grateful to Mrs. Almas Butt, Ms. Shama Khan, Mrs. Shahnaz Zaman (SOS Childrens’ Village, Pakistan), Mrs. Nighat Akbar, and Mr. Muhammad Ali (SOS Hermann Gemeiner Educators School, Pakistan), and Mr. Anwar-ul-Haq (SOS Youth Home, Lahore, Pakistan) for kindly facilitating assessments of PLMS children, control children, and PLMS adults respectively. We are also thankful to Ms. Sumbal Naveed and Ms. Anooshay Abid (Lahore University of Management Sciences, Lahore, Pakistan) for their assistance in data collection; Ms. Niharika Gaur, Ms. Serena Rigotti, Dr. Naguia Haymour, Dr. Alekhya Mazumdar, and Ms. Francesca Manuela (University of Zurich) for assistance in molecular analyses and Sandra Binias (Nencki Institute of Experimental Biology) for bioinformatics support. Finally, we are thankful to the administration and staff of Chughtai Laboratories Private Limited, Pakistan for assistance with blood and seminal fluid collection procedures.

## Authors’ contributions

AJ contributed to study conceptualization, collection of behavioral data, sampling of biofluids, molecular analyses, statistical analyses and visualization, manuscript writing, review and editing, project funding, and project administration and supervision. MG, MK and WT contributed to molecular analyses, statistical analyses and visualization, and assisted in manuscript writing. MM, ZYK, and MT contributed to collection of behavioral data, sampling of biofluids, and data mining. IG, AM, RM, and AC performed molecular analyses in the mouse models. AM and TH assisted in molecular analyses of the human samples. JP performed embryo transplantation. KT performed in vitro experiments and assisted in molecular analyses. AJR and BG performed RNA sequencing of the serum and sperm samples. PF, LL, MF, and NZ performed the metabolomics analyses. SF, OC, and SUC contributed to collection of biofluids and project administration. SG and MP performed the analyses of the omics data. IMM contributed to study conceptualization, manuscript review and editing, project funding, and project administration and supervision. All authors have reviewed and approved the final manuscript.

## EXTENDED DATA MATERIAL

**Extended Data Figure 1.**
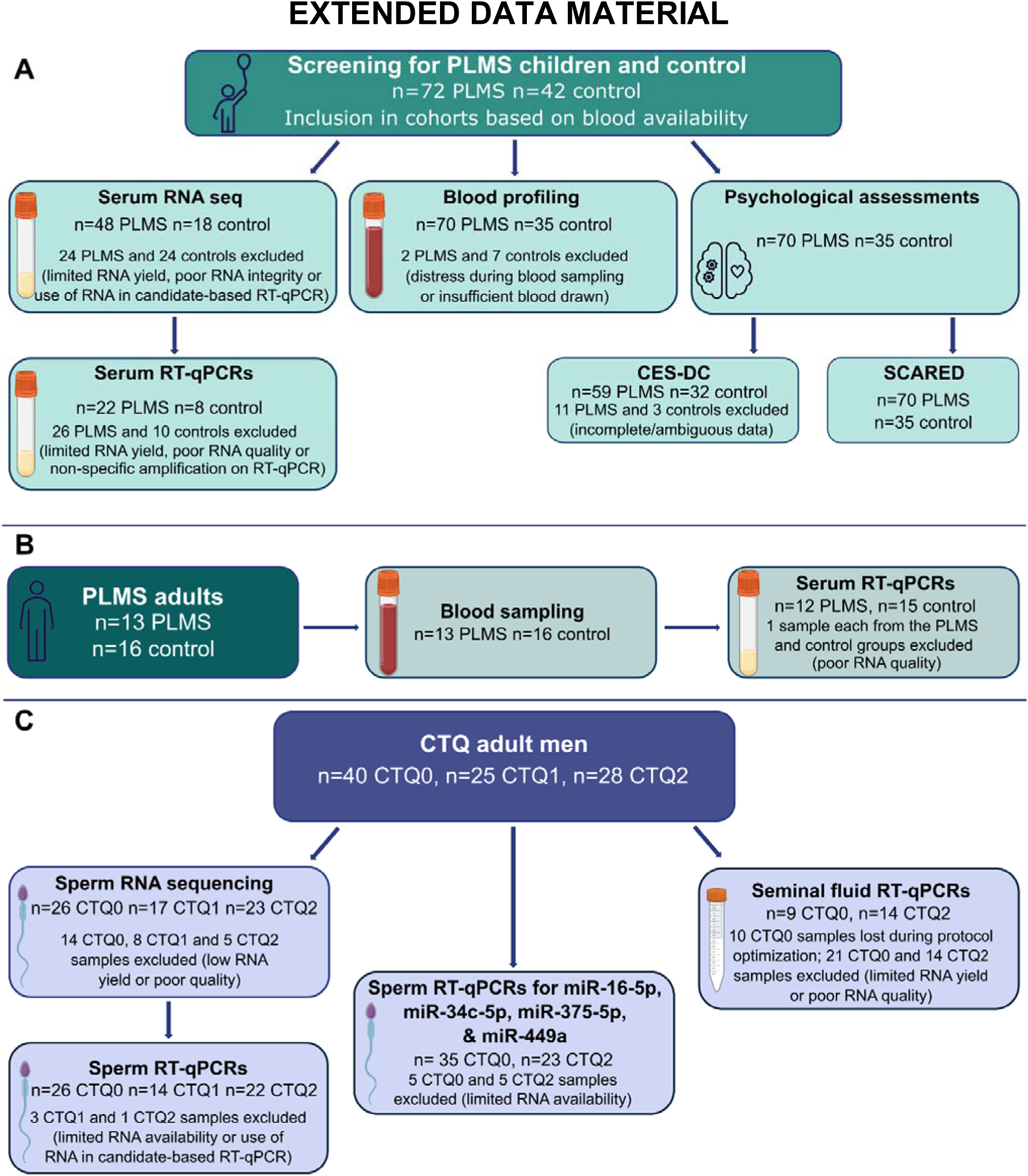
Flow chart of the investigations performed with reasons for variation in sample sizes across **(A)** PLMS children cohort, **(B)** Adult PLMS cohort, and **(C)** CTQ adult cohort.

**Extended Data Figure 2.**
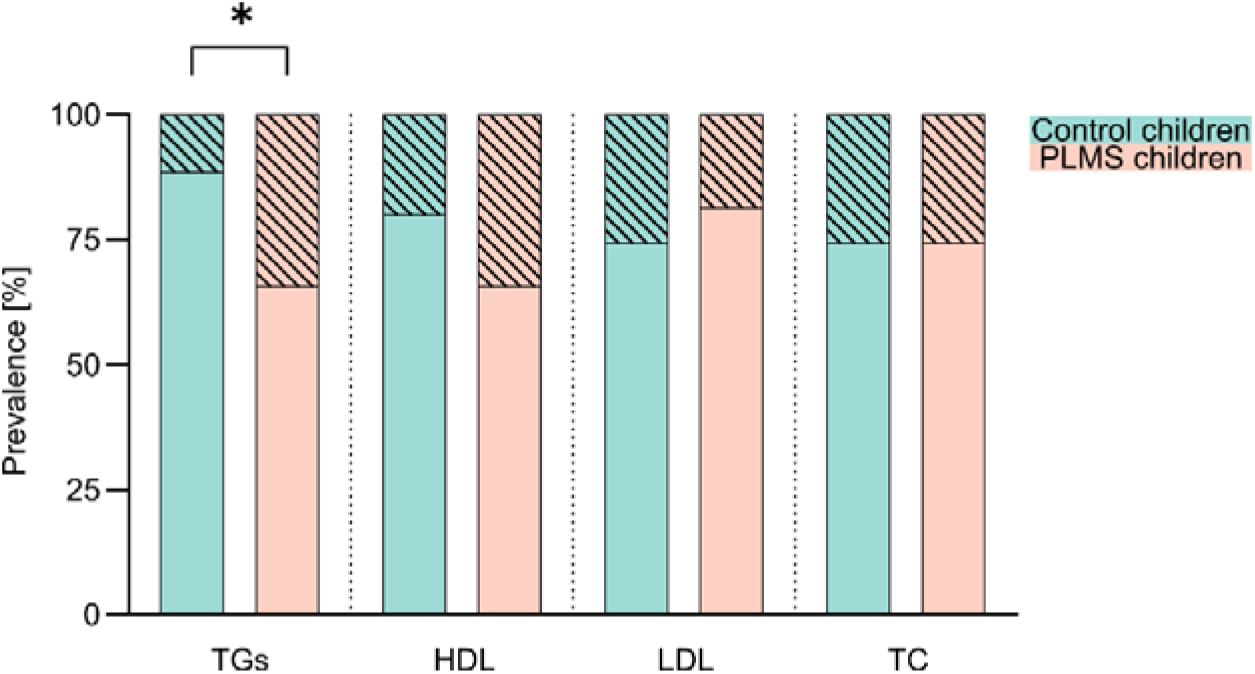
Distribution of non-desirable lipid measures in PLMS (n=70) vs control children (n=35). Non-desirable TG levels: 34% PLMS vs 11% control, *p=0.0181*. Non-desirable HDL levels: 34% PLMS vs 20% control, *p=0.1742*. Non-desirable LDL levels: 19% PLMS vs 26% control, *p=0.4494*. Non-desirable TC levels: 26% PLMS vs 26% control, *p>0.9999*. All analyses were performed using Fisher’s exact test. No outliers were removed. *\* p<0.05*.

**Extended Data Figure 3.**
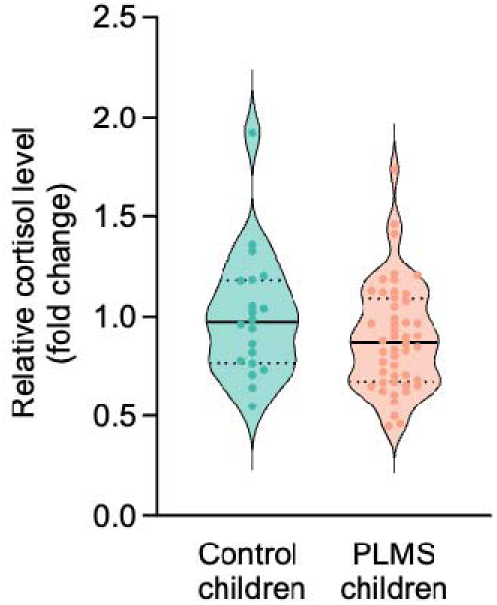
Salivary cortisol in PLMS (n=48) and control (n=20) children. Mean ± SD: 0.89 ± 0.27 PLMS vs 1 ± 0.31 control; unpaired t-test, *p=0.1566*. Outliers were removed using ROUT Q=1%. Violin plots indicate individual values supplemented with group means and IQRs.

**Extended Data Figure 4.**
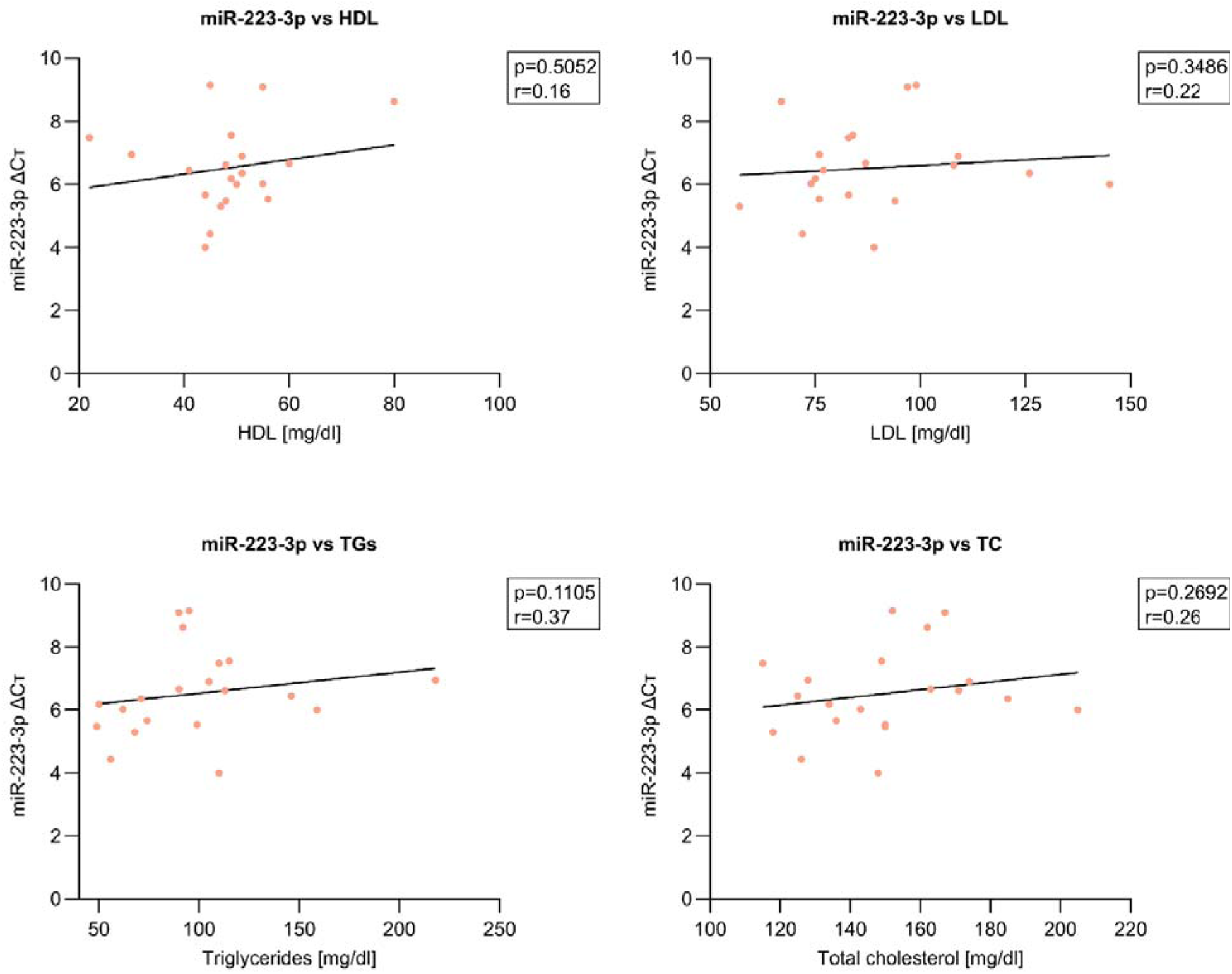
Correlation of serum miR-223-3p level with lipid profiles of PLMS (n=20) children. HDL: *p=0.5052, r=0.16*. LDL: *p=0.3486, r=0.22.* TGs: *p=0.1105, r=0.37.* TC: *p=0.2692, r=0.26.* All analyses were performed using Pearson correlation test.

**Extended Data Figure 5.**
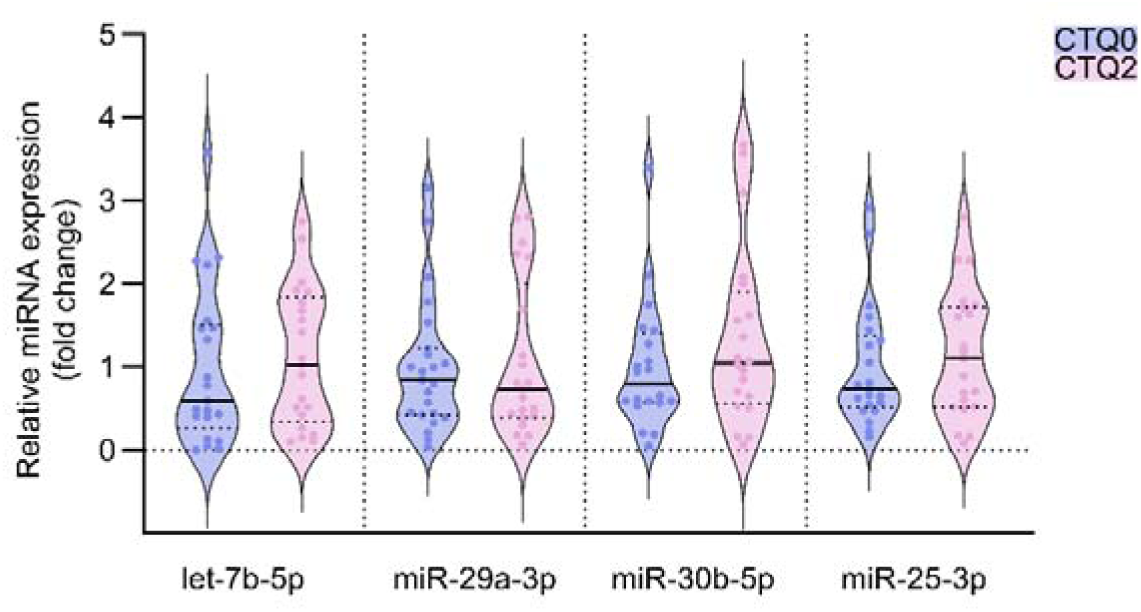
Sperm negative-control miRNAs level in CTQ2 (n=21) and CTQ0 (n=23) men assessed by RT-qPCR. miRNAs were chosen based on the small RNA sequencing. let-7b-5p median [IQR]: 1.03 [0.34–1.84] CTQ2 vs 0.6 [0.27–1.51] CTQ0; *p=0.3963*. miR-29a-3p median [IQR]: 0.73 [0.4–2.01] CTQ2 vs 0.84 [0.43–1.22] CTQ0; *p=0.9444*. miR-30b-5p median [IQR]: 1.05 [0.56–1.9] CTQ2 vs 0.8 [0.57–1.4] CTQ0; *p=0.3547*. miR-25-3p median [IQR]: 1.1 [0.52–1.72] CTQ2 vs 0.74 [0.51–1.38] CTQ0; *p=0.501*. Outliers were removed using ROUT Q=1%. All analyses were performed using Mann-Whitney test. Violin plots indicate individual values supplemented with group means and IQRs.

**Extended Data Figure 6.**
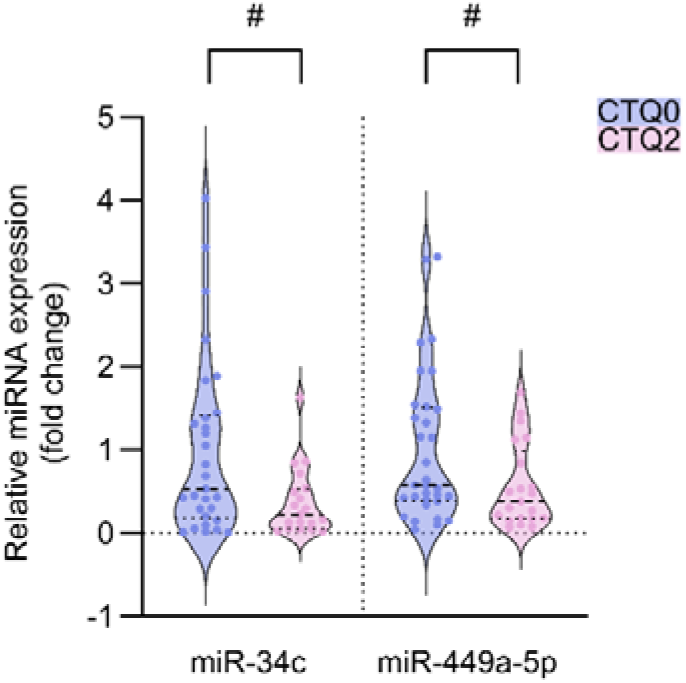
Effects of complex trauma (CTQ2) on the sperm expression of miRNAs previously linked to childhood adversity. Sperm miRNAs level in CTQ2 (n=23) and CTQ0 (n=35) men assessed by RT-qPCR. miR-34c median [IQR]: 0.22 [0.06–0.53] CTQ2 vs 0.53 [0.18–1.41] CTQ0; Mann-Whitney test, *p=0.0518*. miR-449a-5p median [IQR]: 0.38 [0.18–0.98] CTQ2 vs 0.57 [0.39–1.51] CTQ0; Mann-Whitney test, *p=0.0528*. Outliers were removed using ROUT Q=1%. Violin plots indicate individual values supplemented with group means and IQRs. *# p<0.1*.

**Extended Data Figure 7.**
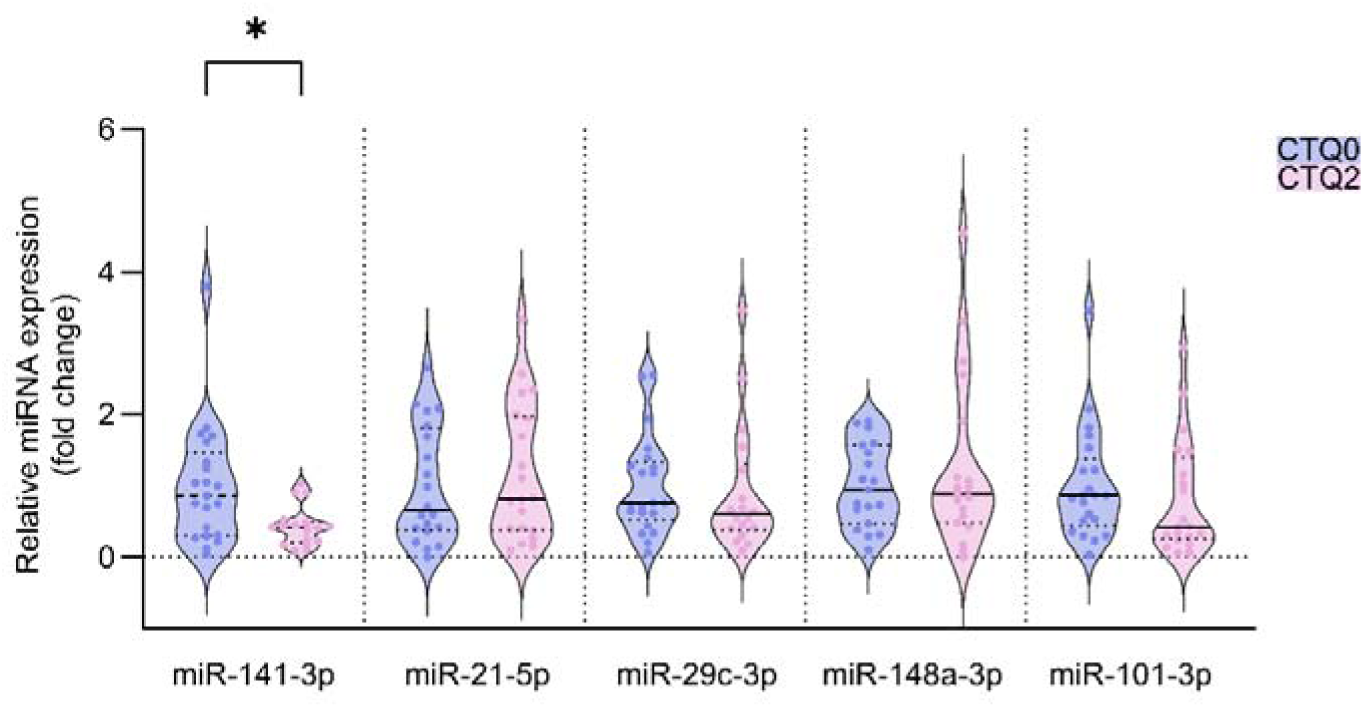
Effects of complex trauma (CTQ2) on the sperm expression of miRNAs linked to childhood adversity in Tuulari et al. Sperm miRNAs level in CTQ2 (n=21) and CTQ0 (n=23) men assessed by RT-qPCR. miR-141-3p median [IQR]: 0.41 [0.2–0.49] CTQ2 vs 0.86 [0.29–1.47] CTQ0; *p=0.0207*. miR-21-5p median [IQR]: 0.81 [0.38–1.98] CTQ2 vs 0.65 [0.37–1.8] CTQ0; *p=0.6205*. miR-29c-3p median [IQR]: 0.6 [0.38–1.3] CTQ2 vs 0.76 [0.53–1.33] CTQ0; *p=0.3351*. miR-148a-3p median [IQR]: 0.89 [0.48–1.9] CTQ2 vs 0.94 [0.47–1.57] CTQ0; *p=0.9310*. miR-101-3p median [IQR]: 0.42 [0.25–1.4] CTQ2 vs 0.86 [0.43–1.37] CTQ0; *p=0.2709*. Outliers were removed using ROUT Q=1%. All analyses were performed using Mann-Whitney test. Violin plots indicate individual values supplemented with group means and IQRs. *\* p<0.05*.

**Extended Data Figure 8.**
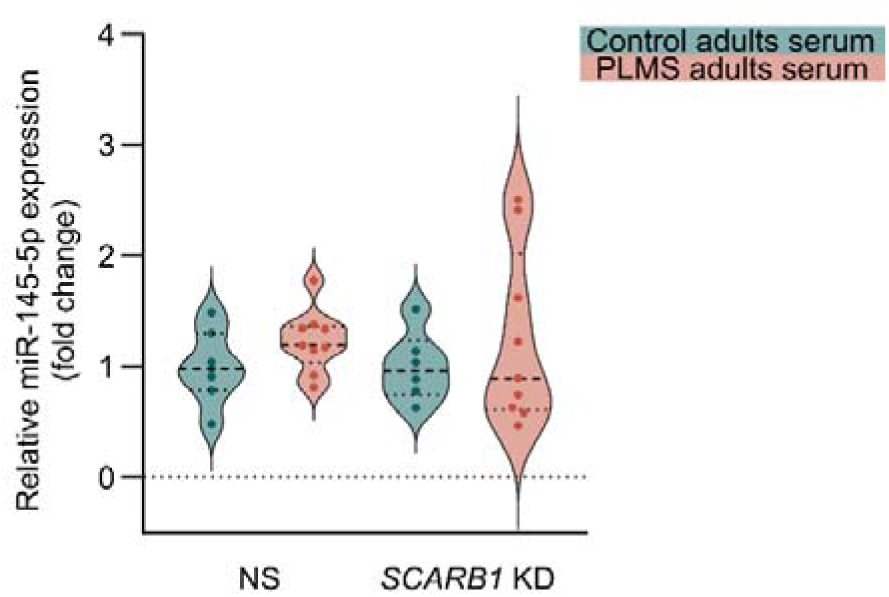
miR-145-5p expression in GC-1 cells transfected with non-silencing control siRNA (NS) or siRNA targeting SR-B1 coding gene (*SCARB1* KD) upon treatment with serum from adult PLMS or control subjects (NS PLMS serum n=9; NS control serum n=7; *SCARB1* KD PLMS serum n=8; *SCARB1* KD control serum n=6). Mean ± SD: 1±0.33 NS (control serum), 1.23±0.28 NS (PLMS serum), 1±0.31 *SCARB1* KD (control serum), 1.23±0.78 (PLMS serum); two-way ANOVA with Tukey’s post-hoc test, *p=0.2083* (PLMS vs control), *p=0.9989* (*SCARB1* KD vs NS), *p=0.9989* (interaction). No outliers were removed. Violin plots indicate individual values supplemented with group means and IQRs.

**Extended Data Figure 9.**
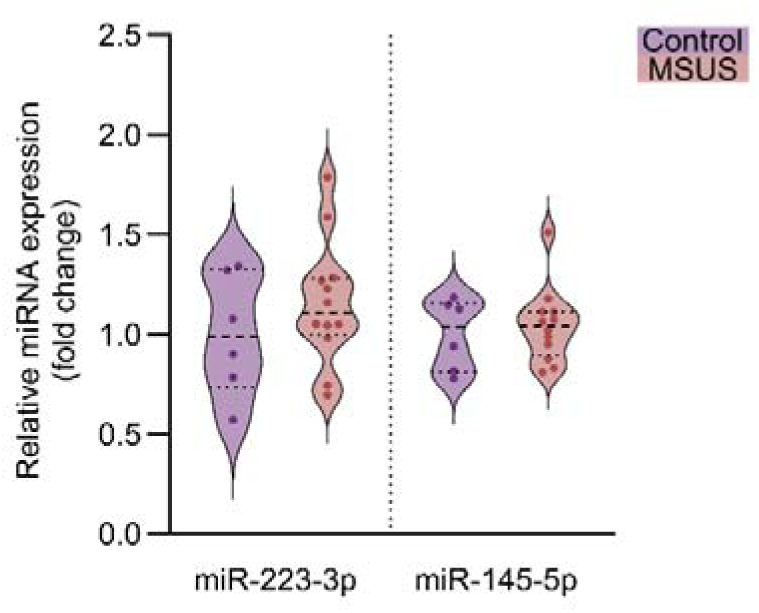
miRNAs levels in the serum of MSUS (n=12) and control (n=6) males assessed by RT-qPCR. miR-223-3p mean ± SD: 1.16 ± 0.31 MSUS vs 1 ± 0.3 control; unpaired t-test, *p=0.3255*. miR-145-5p median [IQR]: 1.04 [0.89–1.11] MSUS vs 1.03 [0.81–1.16] control; Mann-Whitney test, *p=0.9461*. No outliers were removed. Violin plots indicate individual values supplemented with group means and IQRs.

## SUPPLEMENTARY MATERIAL

**Supplementary Table 1.** Demographics of PLMS and control children. Age and hemoglobin comparisons analyzed by unpaired t-test with Welch’s correction. Gender and BMI comparisons analyzed by Chi-square test.

|  | Control children<br>(n = 42) | PLMS children<br>(n = 72)* | p-value |
| --- | --- | --- | --- |
| Age [years] | 9.5 ± 1.8 | 9.9 ± 1.6 | 0.311 |
| Sex |  |  | 0.967 |
| - Females | 20 (47.6%) | 34 (47.2%) |  |
| - Males | 22 (52.4%) | 38 (52.8%) |  |
| BMI categories |  |  | 0.229 |
| - Underweight | 5 (12.8%) | 4 (5.6%) |  |
| - Normal weight | 31 (79.5%) | 61 (84.7%) |  |
| - Overweight | 3 (7.7%) | 7 (9.7%) |  |
| Hemoglobin [g/dl] | 12.63 ± 0.77 | 12.37 ± 1.1 | 0.203 |

\*BMI data unavailable on 3 control children.

**Supplementary Table 2.** Demographics of adult PLMS and adult controls. Age compared by unpaired t-test.

|  | Control adults<br>(n = 16) | PLMS adults<br>(n = 13) | p-value |
| --- | --- | --- | --- |
| Age [years] | 20.5 ± 1.5 | 19.7 ± 1.1 | 0.102 |
| BMI categories |  |  |  |
| - Underweight | 1 (6.25%) | - |  |
| - Normal weight | 9 (56.25%) | 10 (76.9%) |  |
| - Overweight | 6 (37.5%) | 2 (15.4%) |  |
| - Obese | - | 1 (7.7%) |  |

**Supplementary Table 3.**
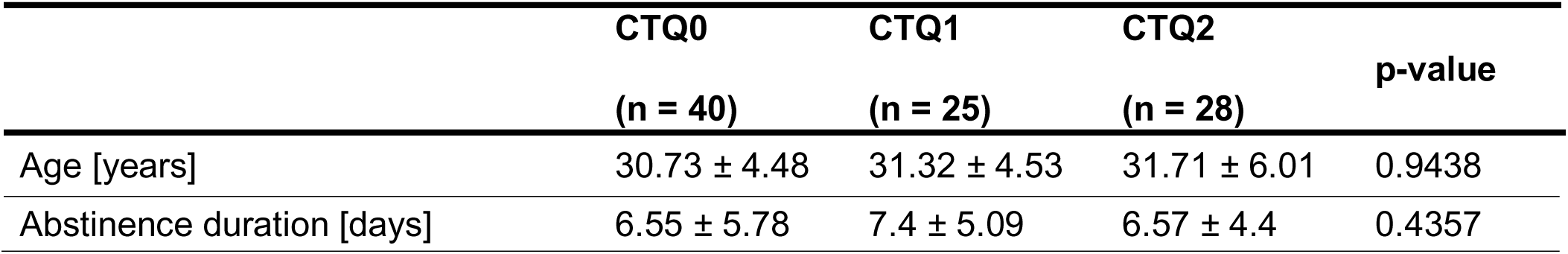
Demographic analysis of participating adults from the CTQ cohort. Age and abstinence duration compared by Kruskal-Wallis test.

**Supplementary Figure 1.**
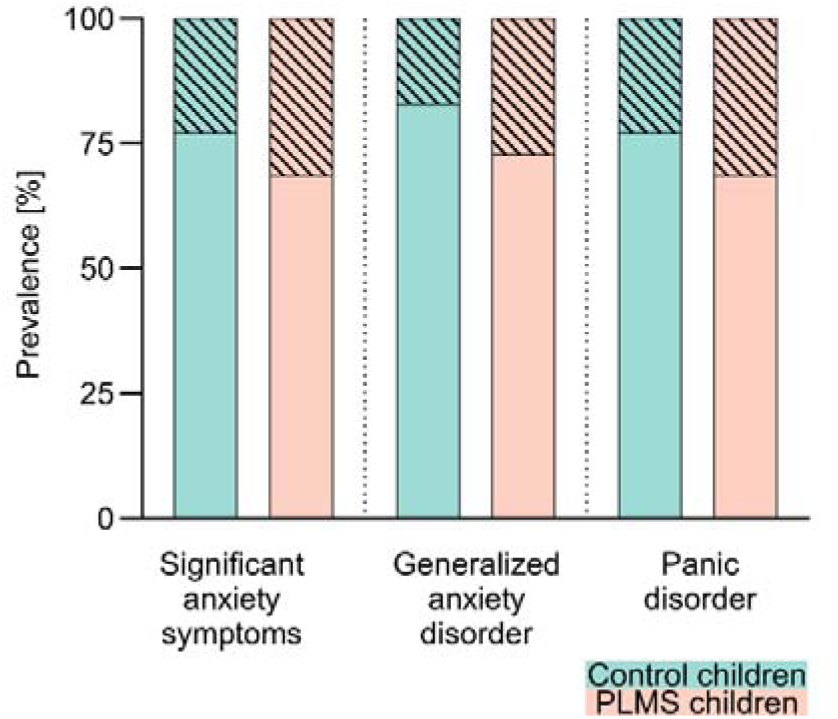
Prevalence [%] of anxiety disorders in PLMS (n=70) and control children (n=35). Significant anxiety symptoms: 31.43% PLMS vs 22.86% control; *p=0.4924*. Generalized anxiety disorder: 27.14% PLMS vs 17.14% control; *p=0.3338*. Panic disorder: 31.43% PLMS vs 22.86% control; *p=0.4924*. No outliers were removed. All analyses were performed using Fisher’s exact test.

**Supplementary Figure 2.**
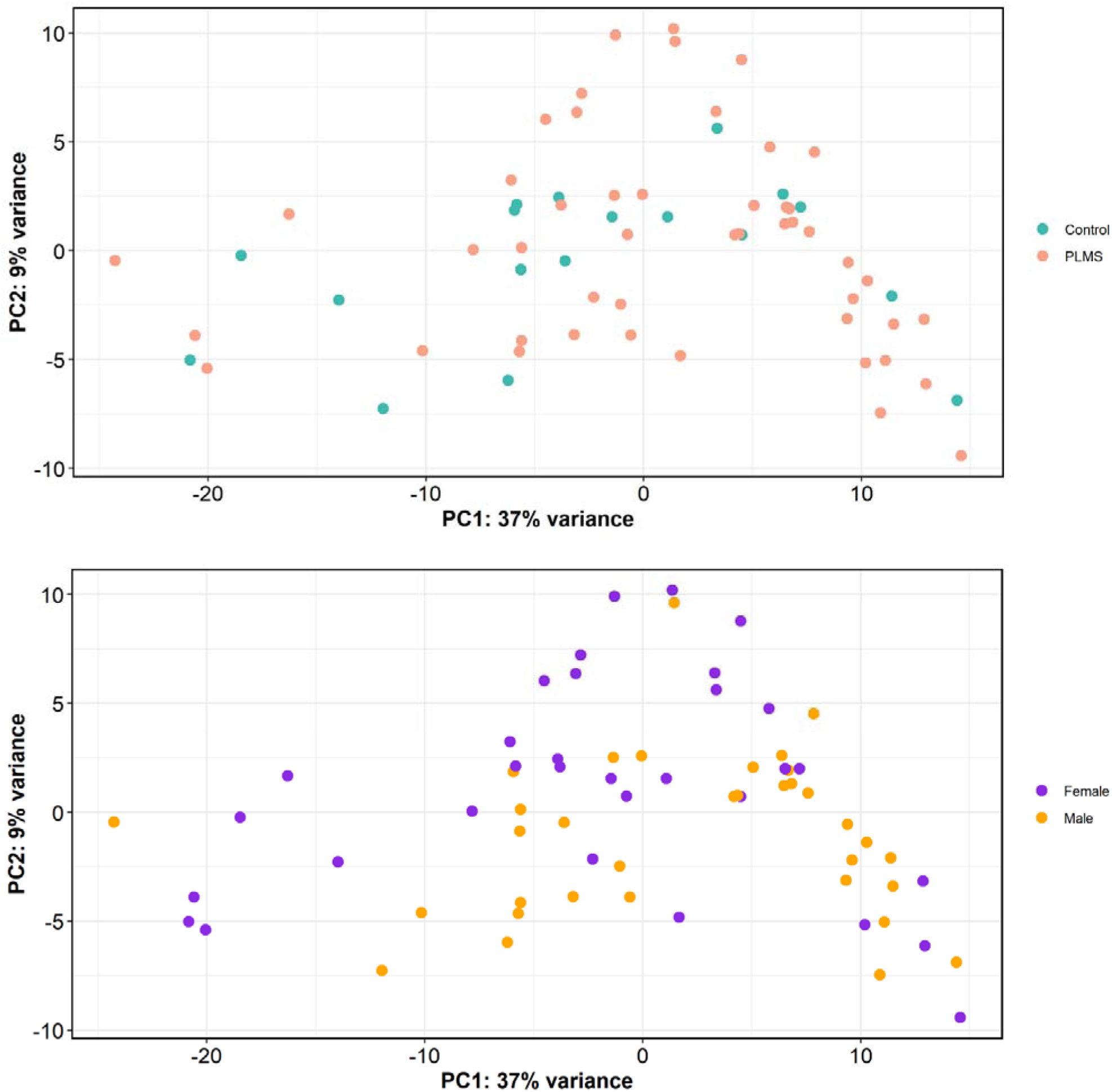
PCA of small RNAs from PLMS (n=48) and control (n=18) children serum. Scatter plots show variance explained by the first two principal components. (Top) PCA of PLMS vs control children. (Bottom) PCA of female vs male children.

**Supplementary Figure 3.**
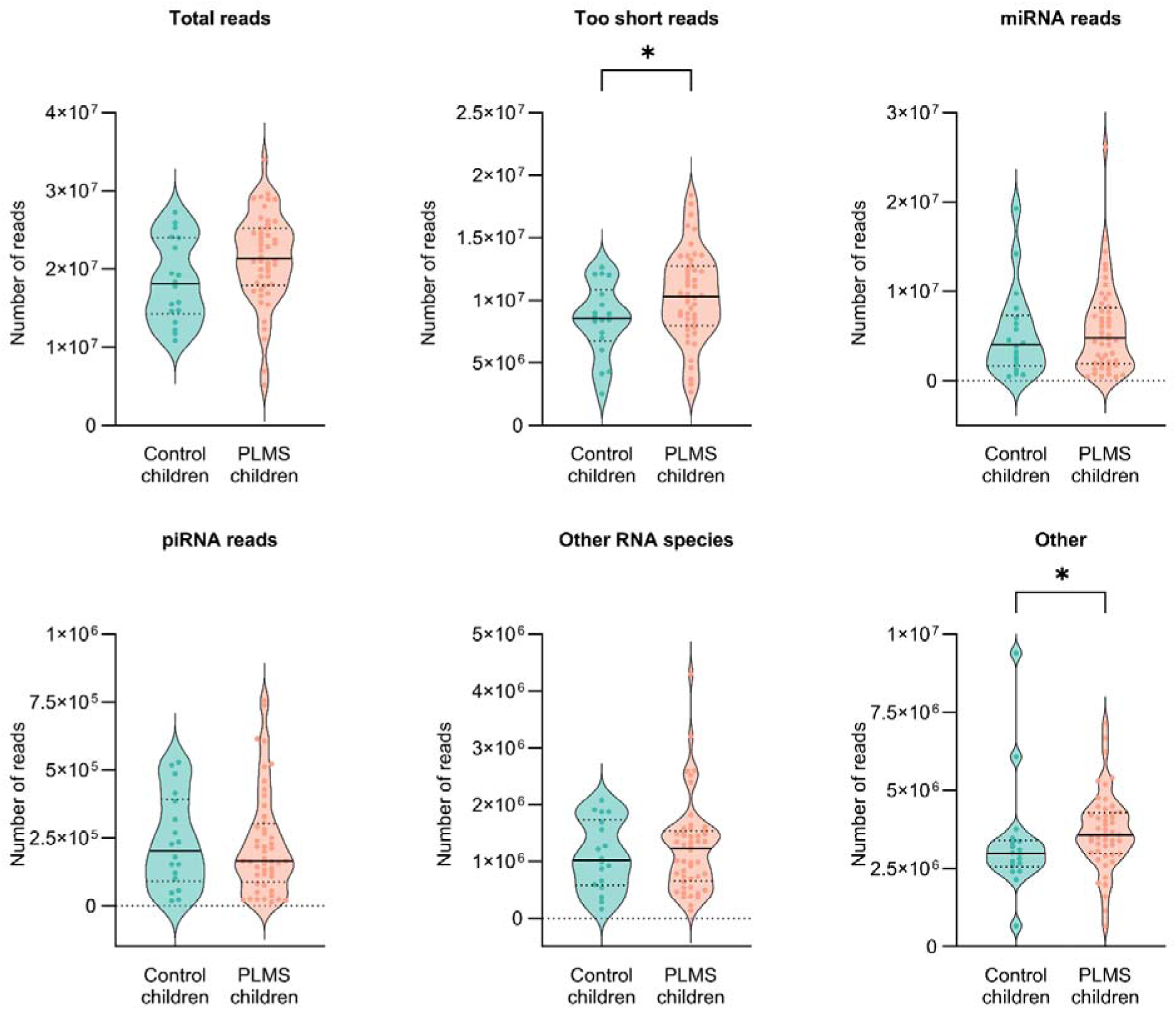
RNA species detected by small RNA sequencing of PLMS (n=48) and control (n=18) children serum. Total reads mean ± SD: 2.1×10^7^ ± 5.8×10^6^ PLMS vs 1.8×10^7^ ± 5.3×10^6^ control; unpaired t-test*, p=0.0727*. Too short reads mean ± SD: 1.0×10^7^ ± 3.6×10^6^ PLMS vs 8.5×10^6^ ± 2.9×10^6^ control; unpaired t-test, *p=0.0480*. miRNA reads median [IQR]: 4.8×10^6^ [1.9×10^6^–8.2×10^6^] PLMS vs 4.0×10^6^ [1.7×10^6^–7.3×10^6^] control; Mann-Whitney test*, p=0.6530*. piRNA reads median [IQR]: 1.6×10^5^ [8.8×10^4^–3.0×10^5^] PLMS vs 2.0×10^5^ [9.0×10^4^–3.9×10^5^] control; Mann-Whitney test, *p=0.7590*. Other RNA species median [IQR]: 1.2×10^6^ [6.5×10^5^–1.5×10^6^] PLMS vs 1.0×10^6^ [5.8×10^5^–1.7×10^6^] control; Mann-Whitney test, *p=0.9250*. Other median [IQR]: 3.6×10^6^ [3.0×10^6^–4.3×10^6^] PLMS vs 3.0×10^6^ [2.5×10^6^–3.4×10^6^]; Mann-Whitney test, *p=0.0233*. No outliers were removed. Violin plots indicate individual values supplemented with group means and IQRs. *\* p<0.05*.

**Supplementary Figure 4.**
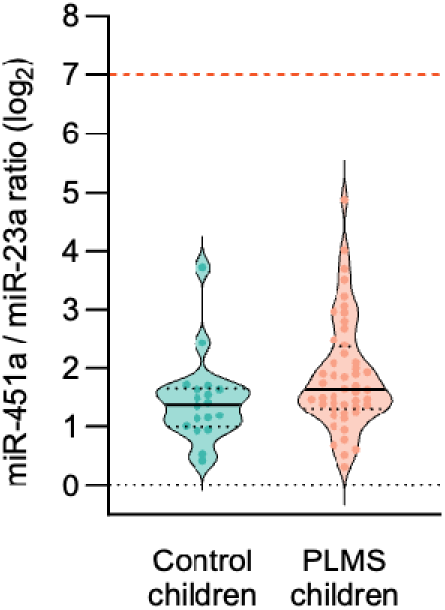
Hemolysis control - miR-451 to miR-23a ratio based on log2 TMM (trimmed mean of values)-normalized counts per million from small RNA sequencing of PLMS (n=48) and control (n=18) children serum. Ratio >7 indicates significant hemolysis and sample compromise.

**Supplementary Figure 5.**
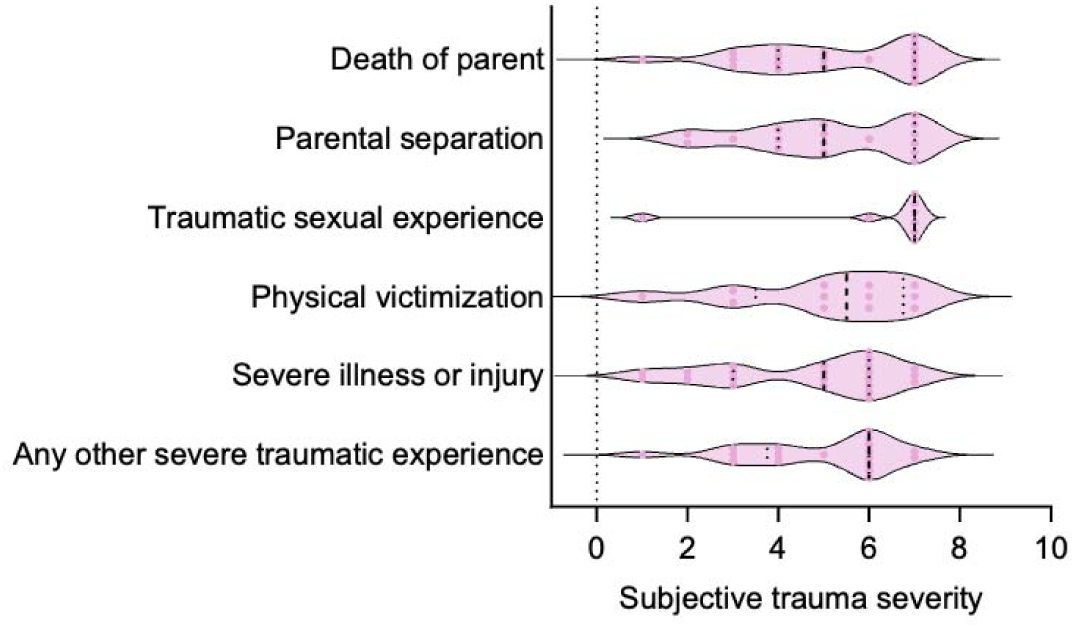
Severity of different trauma types assessed by CTQ in the adult men cohort (cohort n=93, death of parent n=20, parental separation n=17, traumatic sexual experience n=8, physical victimization n=12, severe illness or injury n=23, any other severe traumatic experience n=18). Non-significant (*p=0.7825*) difference according to one-way ANOVA.

**Supplementary Figure 6.**
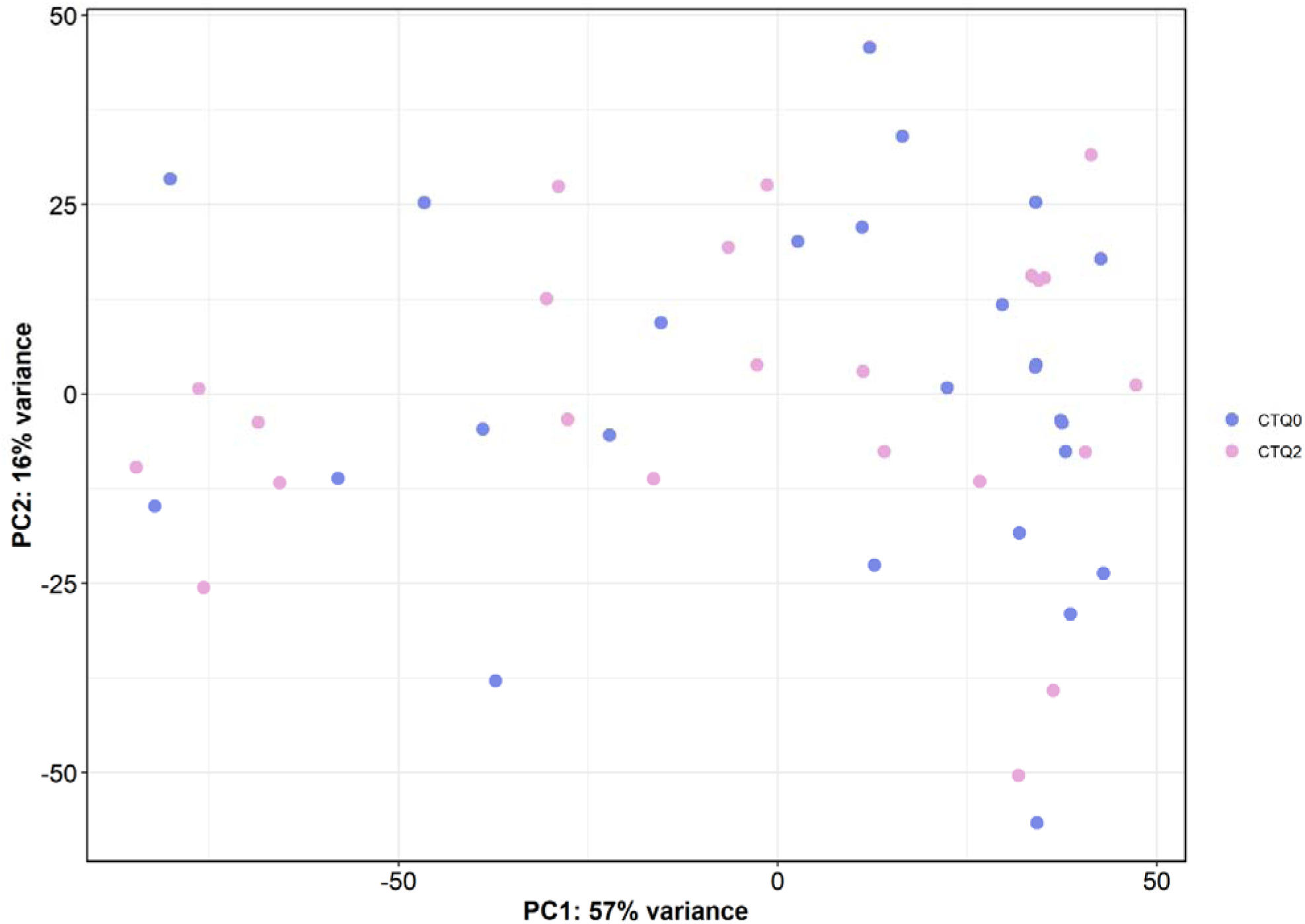
PCA of small RNAs from CTQ2 (n=23) vs CTQ0 (n=26) men. Scatter plot shows variance explained by the first two principal components.

**Supplementary Figure 7.**
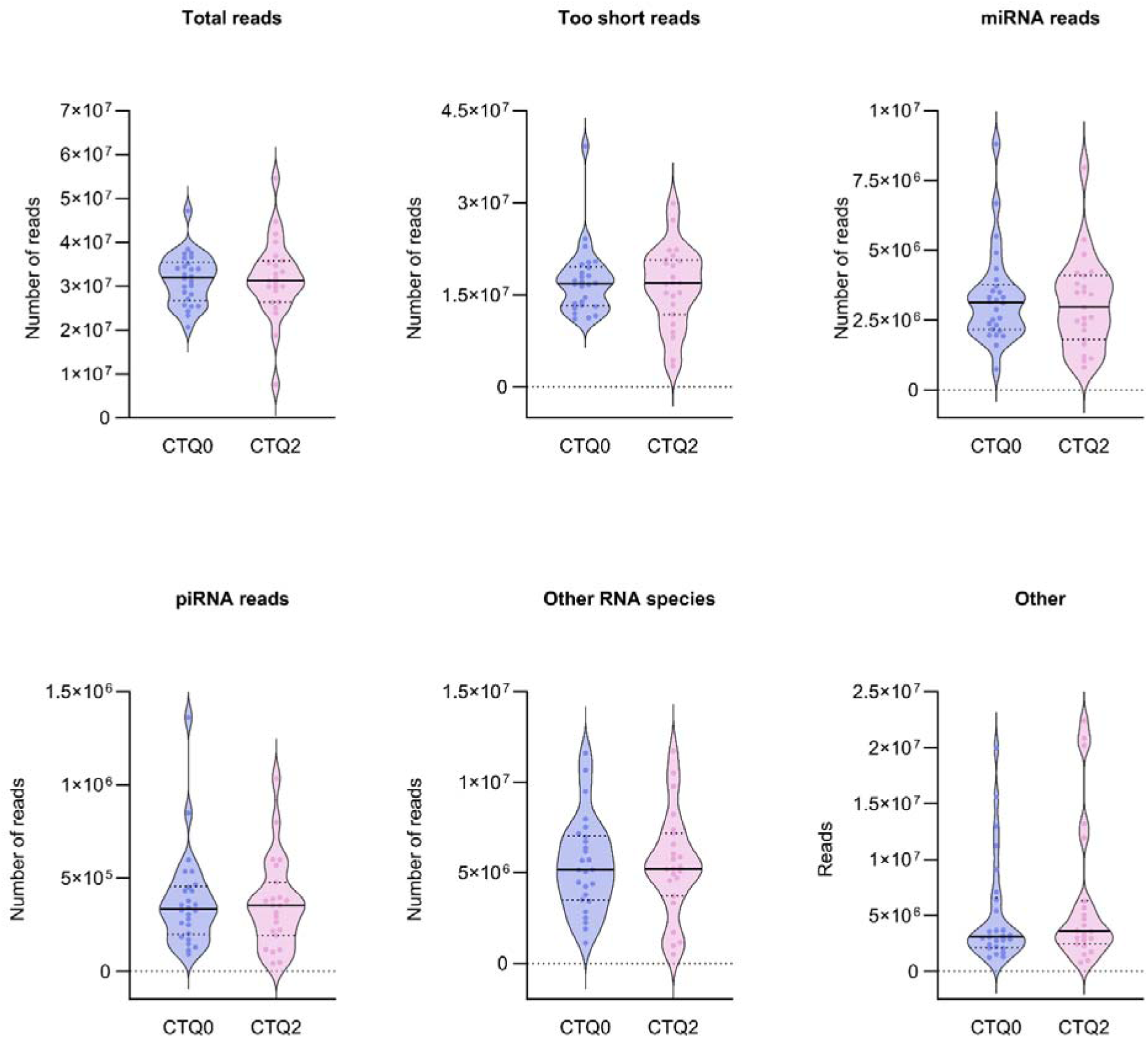
RNA species detected by small RNA sequencing of CTQ2 (n=23) and CTQ0 (n=26). Total reads mean ± SD: 3.2×10^7^ ± 9.2×10^6^ CTQ2 vs 3.2×10^7^ ± 5.8×10^6^ CTQ0; unpaired t-test Welch’s correction, *p=0.9408*. Too short reads median [IQR]: 1.7×10^7^ [1.2×10^7^–2.1×10^7^] CTQ2 vs 1.7×10^7^ [1.3×10^7^–2.0×10^7^] CTQ0; Mann-Whitney test, *p=0.9446*. miRNA reads median [IQR]: 2.9×10^6^ [1.8×10^6^–4.1×10^6^] CTQ2 vs 3.1×10^6^ [2.2×10^6^–3.8×10^6^] CTQ0; Mann-Whitney test, *p=0.8349*. piRNA reads median [IQR]: 3.5×10^5^ [1.9×10^5^–4.8×10^5^] CTQ2 vs 3.3×10^5^ [2.0×10^5^–4.5×10^5^] CTQ0; Mann-Whitney test, *p=0.7590*. Other RNA species mean ± SD: 5.4×10^6^ ± 2.9×10^6^ CTQ2 vs 5.4×10^6^ ± 2.6×10^6^ CTQ0; unpaired t-test; p=*0.9995*. Other Median [IQR]: 3.6×10^6^ [2.4×10^6^–6.3×10^6^] CTQ2 vs 3.1×10^6^ [2.1×10^6^–6.6×10^6^] CTQ0; Mann-Whitney test, *p=0.5713*. No outliers were removed. Violin plots indicate individual values supplemented with group means and IQRs.

**Supplementary Figure 8.**
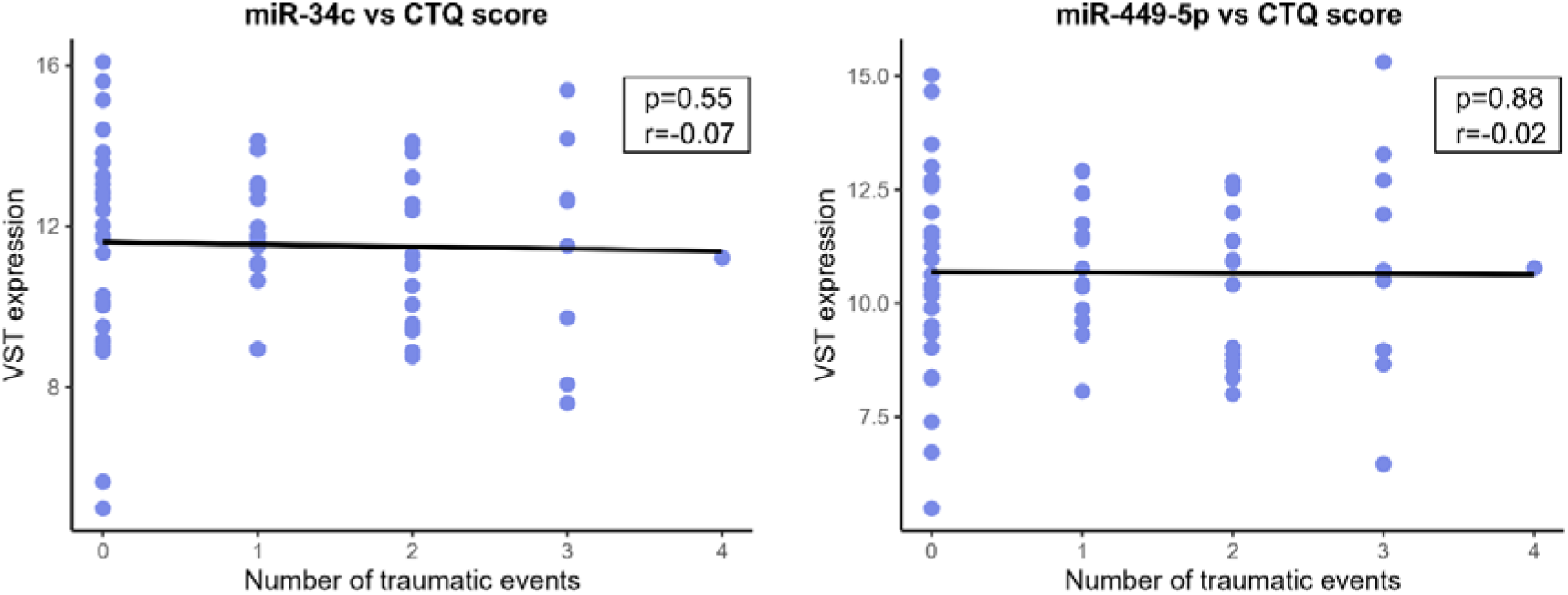
Correlation of sequencing-based sperm miRNAs level with the number of traumatic events (n=66). miR-34c: Pearson correlation, *p=0.55, r=-0.07*. miR-449a-5p: Pearson correlation, *p=0.88, r=-0.02*. VST (variance-stabilizing

**Supplementary Figure 9.**
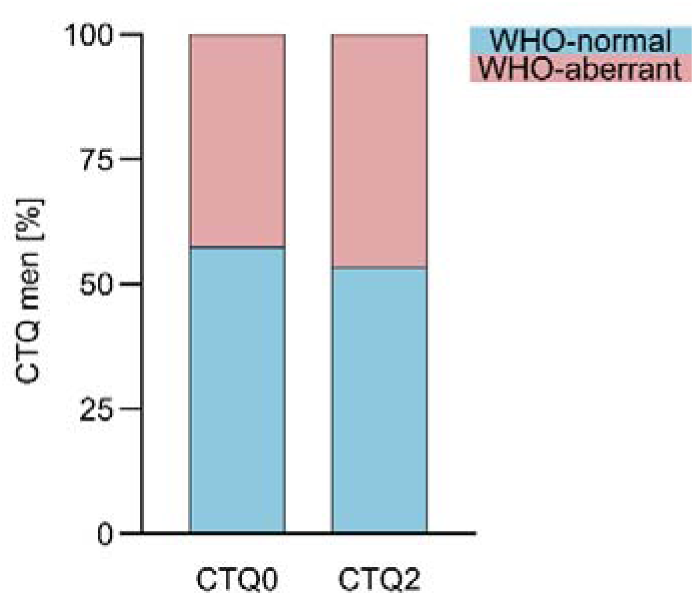
The proportion of normal vs aberrant sperm in the samples from CTQ2 (n=28) vs CTQ0 (n=40) subjects according to the WHO criteria. CTQ2 aberrant = 46.43% vs CTQ0 aberrant = 42.5%; Fisher’s exact test, *p=0.8071*.

**Supplementary Figure 10.**
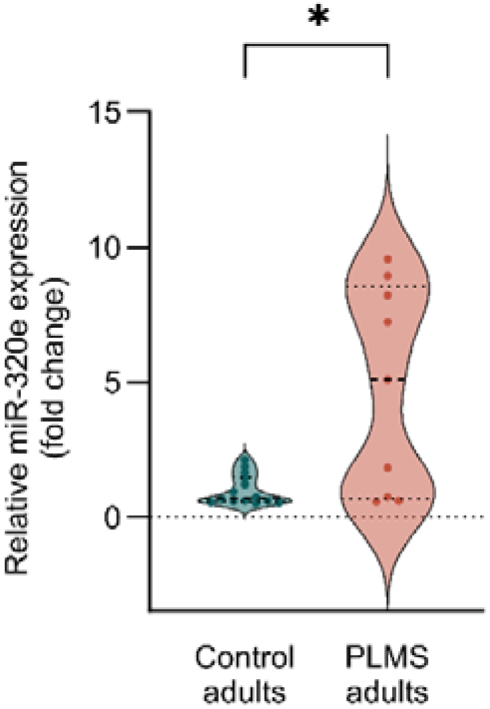
Serum miR-320e level in PLMS (n=9) and control (n=15) adults assessed by RT-qPCR assays. Median [IQR]: 5.08 [0.67–8.57] PLMS vs 0.71 [0.58–1.49] control; Mann-Whitney test, *p=0.0240*. Outliers were removed using ROUT Q=1%. Violin plots indicate individual values supplemented with group means and IQRs. *\* p<0.05*.

**Supplementary Figure 11.**
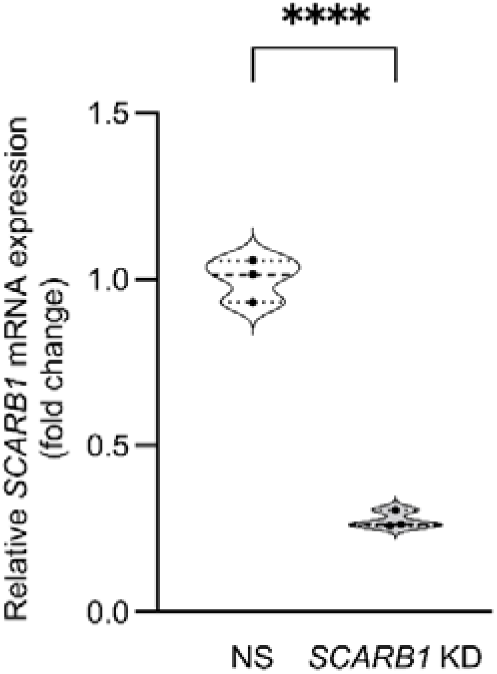
Confirmation of *SCARB1* knockdown in GC1 cells. *SCARB1* mRNA level in *SCARB1* KD (n=3) and NS group (n=3) assessed by RT-qPCR assays. Mean ± SD0.28 ± 0.03 *SCARB1* KD vs 1 ± 0.06 control RNA; unpaired t-test, *p<0.0001*. Violin plots indicate individual values supplemented with group means and IQRs. *\*\*\*\* p<0.0001*.

**Supplementary Figure 12.**
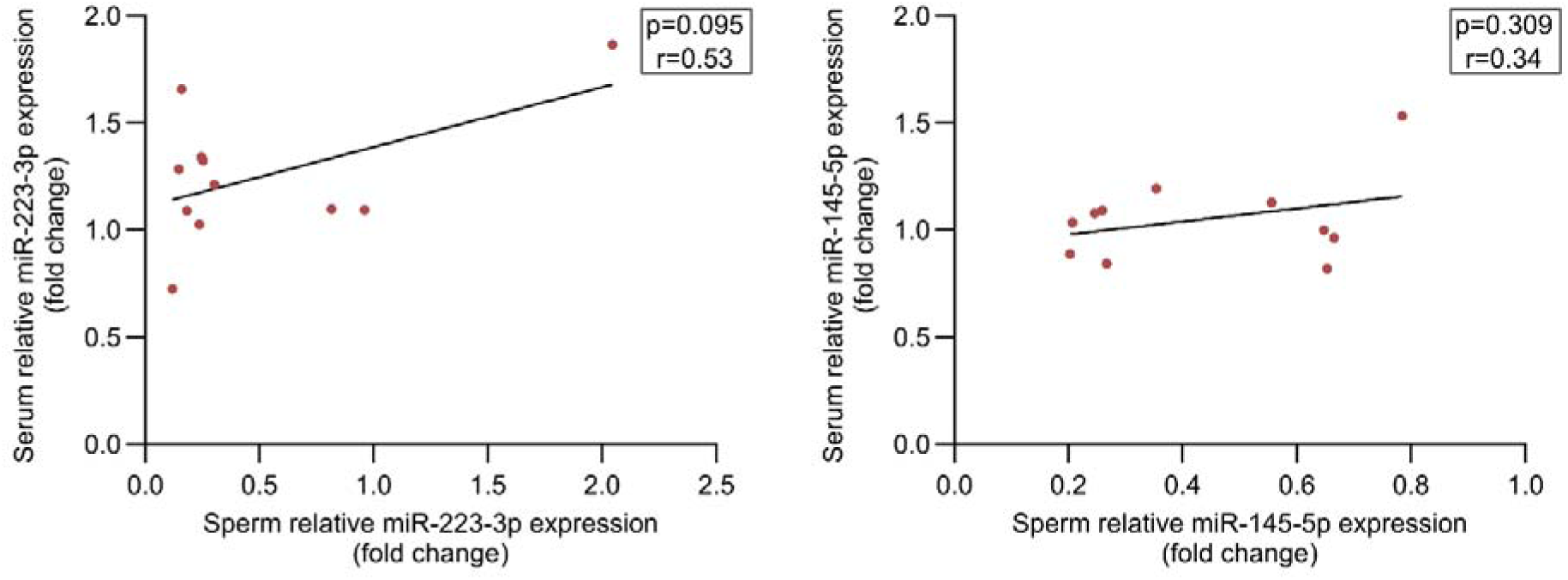
Correlation of serum and sperm miRNAs level in MSUS male mice (n=11). miR-223-3p: Pearson correlation, *p=0.0953, r=0.53*. miR-145-5p: Pearson correlation, *p=0.3088, r=0.34*.

**Supplementary Figure 13.**
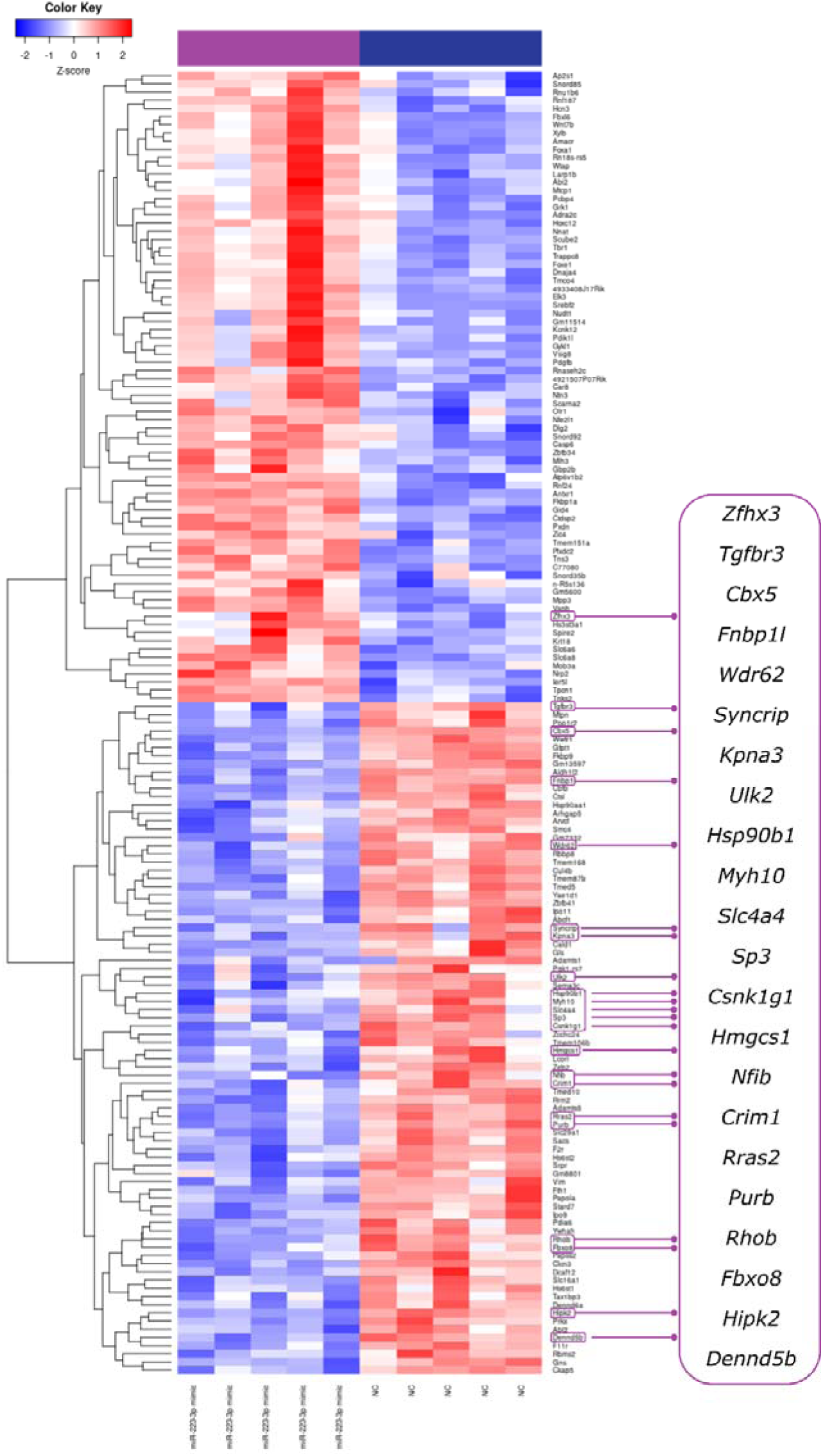
Differential expression of coding genes in GC-1 cells following transfection with miR-223-3p mimic. Heatmap of differentially expressed coding genes with *p.adj. <0.05*. miR-223-3p mimic (n=5), negative control (NC n=5). Inlet shows miR-223-3p targets based on TargetScanHuman database with downregulation of all targets in the miR-223-3p mimic-treated GC-1 cells with the exception of *Zhfx3*.

