## Supplementary statistics for "Differential microRNAs in human serum and sperm after childhood trauma with potential implications for offspring health"

**Supplementary Statistics**. Data are presented as mean ± SD or median (IQR), as appropriate. Exact p-values are reported.

| **Figure** | **Panel** | **Comparison** | **Test** | **n (after ROUT if applicable)** | **Statistic** | **p-value** | **Data summary** |
| --- | --- | --- | --- | --- | --- | --- | --- |
| Fig.1 | A | Control vs PLMS (children) | Welch’s t-test | 32/59 | t = 2.46; df = 84.14 | 0.0161 | Control: 12.63 ± 2.95; PLMS: 15.27 ± 7.25 |
| Fig.1 | B | Control vs PLMS (children) | Fisher’s exact test | 32/59 | OR = 3.01 | **0.0401** | Control: 25/7; PLMS: 32/27 |
| Fig.1 | C | Control vs PLMS (children) | Mann-Whitney test | 35/69 | U = 861 | **0.016** | Control: 7 (5–8); PLMS: 8 (5–12.5) |
| Fig.1 | D (TGs) | Control vs PLMS (children) | Mann-Whitney test | 35/70 | U = 748 | **0.001** | Control: 79 (60–100); PLMS: 106 (76.5–145.3) |
| Fig.1 | D (HDL) | Control vs PLMS (children) | Unpaired t-test | 35/70 | t = 0.86; df = 103 | 0.3898 | Control: 50.63 ± 7.99; PLMS: 48.87 ± 10.62 |
| Fig.1 | D (LDL) | Control vs PLMS (children) | Mann-Whitney test | 35/70 | U = 1001 | 0.1278 | Control: 91 (82–113); PLMS: 84 (74–105) |
| Fig.1 | D (TC) | Control vs PLMS (children) | Mann-Whitney test | 35/70 | U = 1082 | 0.3334 | Control: 154 (140–171); PLMS: 150 (133.8–170.3) |
| Fig.2 | C (miR-223-3p) | Control vs PLMS (children) | Mann-Whitney test | 7/21 | U = 24 | **0.007** | Control: 0.66 (0.21–1.27); PLMS: 2.92 (1.39–4.5) |
| Fig.2 | C (miR-144-3p) | Control vs PLMS (children) | Mann-Whitney test | 7/19 | U = 19 | **0.0045** | Control: 1.3 (0.3–1.63); PLMS: 4.81 (1.51–9.15) |
| Fig.2 | D (miR-223-3p) | Control vs PLMS (adults) | Mann-Whitney test | 15/11 | U = 34 | **0.0108** | Control: 0.91 (0.55–1.18); PLMS: 2.66 (0.83–2.76) |
| Fig.2 | D (miR-144-3p) | Control vs PLMS (adults) | Welch’s t-test | 14/12 | t = 3.34; df = 11.79 | **0.006** | Control: 1 ± 0.58; PLMS: 3.77 ± 2.83 |
| Fig.3 | C (miR-223-3p) | CTQ0 vs CTQ2 (sperm) | Mann-Whitney test | 21/18 | U = 100 | **0.0188** | CTQ0: 0.44 (0.12–1.19); CTQ2: 1.52 (0.48–6.81) |
| Fig.3 | C (miR-145-5p) | CTQ0 vs CTQ2 (sperm) | Mann-Whitney U test | 15/13 | U = 37 | **0.0044** | CTQ0: 0.89 (0.2–1.51); CTQ2: 2.54 (0.99–4.83) |
| Fig.3 | D (miR-223-3p) | Number of CT events vs ΔCT | Pearson correlation | 62 | r = -0.25, 95% CI [-0.47, -0.005]; df = 60 | **0.046** |  |
| Fig.3 | D (miR-145-5p) | Number of CT events vs ΔCT | Pearson correlation | 40 | r = -0.25, 95% CI [-0.52, -0.07]; df = 38 | 0.1277 |  |
| Fig.3 | E (miR-223-3p) | WHO-normal vs WHO-aberrant | Mann-Whitney test | 20/18 | U = 142 | 0.2765 | WHO-normal: 0.3 (0.12–1.55); WHO-aberrant: 0.66 (0.22–1.25) |
| Fig.3 | E (miR-145-5p) | WHO-normal vs WHO-aberrant | Mann-Whitney test | 17/12 | U = 92 | 0.6788 | WHO-normal: 0.87 (0.2–1.52); WHO-aberrant: 0.4 (0.14–1.9) |
| Fig.4 | A (miR-223-3p) | CTQ0 vs CTQ2 (seminal fluid) | Mann-Whitney test | 13/8 | U = 50 | 0.9155 | CTQ0: 0.89 (0.09–1.54); CTQ2: 0.53 (0.27–6.99) |
| Fig.4 | A (miR-145-5p) | CTQ0 vs CTQ2 (seminal fluid) | Mann-Whitney test | 12/7 | U = 21 | 0.0831 | CTQ0: 0.79 (0.52–1.42); CTQ2: 2.83 (0.88–4.36) |
| Fig.4 | B | Control vs PLMS (adults) | Welch’s t-test | 15/9 | t = 2.54; df = 8.73 | **0.0325** | Control: 1.0 ± 0.67; PLMS: 3.1 ± 2.43 |
| Fig.4 | D | Cell line, type of serum and interaction | Two-way ANOVA + Tukey’s test | 6–9/ group | Interaction: F = 0.64; DF = 1.25  Cell line: F = 39.59; DF = 1.25  Serum: F = 0.0043; DF = 1.25 | Interaction: 0.4322; Cell line: **<0.0001**; Serum: 0.9485 | Tukey: NS vs SRB1 KD (all comparisons p ≤ 0.0054) |
| Fig.5 | B (miR-223-3p) | Control vs MSUS (sperm) | Welch’s t-test | 7/9 | t = 4.75; df = 6.14 | **0.003** | Control: 1 ± 0.45; MSUS: 0.18 ± 0.06 |
| Fig.5 | B (miR-145-5p) | Control vs MSUS (sperm) | Mann-Whitney test | 7/11 | U = 9 | **0.0059** | Control: 1.21 (0.46–1.44); MSUS: 0.23 (0.2–0.55) |
| Fig.5 | C (miR-16-5p) | CTQ0 vs CTQ2 | Mann-Whitney test | 35/21 | U = 247 | **0.0411** | CTQ0: 0.8 (0.52–1.51); CTQ2: 0.53 (0.32–1.15) |
| Fig.5 | C (miR-375-5p) | CTQ0 vs CTQ2 | Mann-Whitney test | 33/21 | U = 198 | **0.0077** | CTQ0: 0.81 (0.52–1.47); CTQ2: 0.38 (0.25–0.68) |
| Fig.6 | B (brain) | NC vs miR-223-3p mimic | Unpaired t-test | 4/6 | t = 2.7; df = 8 | **0.0269** | NC: 1 ± 0.08; miR-223-3p mimic: 1.22 ± 0.14 |
| Fig.6 | B (WAT) | NC vs miR-223-3p mimic | Unpaired t-test | 6/6 | t = 20.4; df = 10 | **<0.0001** | NC: 1 ± 0.05; miR-223-3p mimic: 1.9 ± 0.1 |
| Fig.6 | B (BAT) | NC vs miR-223-3p mimic | Unpaired t-test | 6/6 | t = 1.69; df = 10 | 0.1220 | NC: 1 ± 0.42; miR-223-3p mimic: 1.8 ± 1.1 |
| Fig.6 | C (*Htr1a*) | NC vs miR-223-3p mimic | Unpaired t-test | 4/6 | t = 28.11; df = 8 | **<0.0001** | NC: 1 ± 0.11; miR-223-3p mimic: 6.21 ± 0.35 |
| Fig.6 | C (*Htr2a*) | NC vs miR-223-3p mimic | Unpaired t-test | 4/6 | t = 10.59; df = 8 | **<0.0001** | NC: 1 ± 0.1; miR-223-3p mimic: 0.5 ± 0.04 |
| Fig.6 | C (*Creb1*) | NC vs miR-223-3p mimic | Unpaired t-test | 4/6 | t = 4.98; df = 8 | **0.0011** | NC: 1 ± 0.26; miR-223-3p mimic: 1.61 ± 0.13 |
| Ext. Data Fig.2 | TGs | Control vs PLMS (children) | Fisher’s exact test | 35/70 | OR = 4.04 | **0.0181** | Control: 31/4; PLMS 46/24 |
| Ext. Data Fig.2 | HDL | Control vs PLMS (children) | Fisher’s exact test | 35/70 | OR = 2.09 | 0.1742 | Control: 28/7; PLMS 46/24 |
| Ext. Data Fig.2 | LDL | Control vs PLMS (children) | Fisher’s exact test | 35/70 | OR = 0.66 | 0.4494 | Control: 26/9; PLMS: 57/13 |
| Ext. Data Fig.2 | TC | Control vs PLMS (children) | Fisher’s exact test | 35/70 | OR = 1 | >0.9999 | Control: 26/9; PLMS: 52/18 |
| Ext. Data Fig.3 | Cortisol | Control vs PLMS (children) | Unpaired t-test | 20/48 | t = 1.43; df = 66 | 0.1566 | Control: 1 ± 0.31; PLMS: 0.89 ± 0.27 |
| Ext. Data Fig.4 | Upper left | HDL vs ΔCT | Spearman correlation | 20 | r = 0.16, 95% CI [-0.32, 0.57]; df = 18 | 0.5052 |  |
| Ext. Data Fig.4 | Upper right | LDL vs ΔCT | Spearman correlation | 20 | r = 0.22, 95% CI [-0.26, 0.61]; df = 18 | 0.3486 |  |
| Ext. Data Fig.4 | Bottom left | TGs vs ΔCT | Spearman correlation | 20 | r = 0.37, 95% CI [-0.1, 0.7]; df = 18 | 0.1105 |  |
| Ext. Data Fig.4 | Bottom right | TC vs ΔCT | Spearman correlation | 20 | r = 0.26, 95% CI [-0.22, 0.64]; df = 18 | 0.2692 |  |
| Ext. Data Fig.5 | let-7b-5p | CTQ0 vs CTQ2 | Mann-Whitney test | 21/21 | U = 186 | 0.3963 | CTQ0: 0.6 [0.27–1.51]; CTQ2: 1.03 [0.34–1.84] |
| Ext. Data Fig.5 | miR-29a-3p | CTQ0 vs CTQ2 | Mann-Whitney test | 23/21 | U = 238 | 0.9444 | CTQ0: 0.84 [0.43–1.22]; CTQ2: 0.73 [0.4–2.01] |
| Ext. Data Fig.5 | miR-30b-5p | CTQ0 vs CTQ2 | Mann-Whitney test | 20/20 | U = 165 | 0.3547 | CTQ0: 0.8 [0.57–1.4]; CTQ2: 1.05 [0.56–1.9] |
| Ext. Data Fig.5 | miR-25-3p | CTQ0 vs CTQ2 | Mann-Whitney test | 21/21 | U = 193 | 0.501 | CTQ0: 0.74 [0.51–1.38]; CTQ2: 1.1 [0.52–1.72] |
| Ext. Data Fig.6 | miR-34c | CTQ0 vs CTQ2 | Mann-Whitney test | 29/19 | U = 183 | 0.0518 | CTQ0: 0.53 [0.18–1.41]; CTQ2: 0.22 [0.06–0.53] |
| Ext. Data Fig.6 | miR-449a-5p | CTQ0 vs CTQ2 | Mann-Whitney test | 32/21 | U = 229.5 | 0.0528 | CTQ0: 0.57 [0.39–1.51]; CTQ2: 0.38 [0.18–0.98] |
| Ext. Data Fig.7 | miR-141-3p | CTQ0 vs CTQ2 | Mann-Whitney test | 21/17 | U = 100 | **0.0207** | CTQ0: 0.86 [0.29–1.47]; CTQ2: 0.41 [0.2–0.49] |
| Ext. Data Fig.7 | miR-21-5p | CTQ0 vs CTQ2 | Mann-Whitney test | 20/20 | U = 181 | 0.6205 | CTQ0: 0.65 [0.37–1.8]; CTQ2: 0.81 [0.38–1.98] |
| Ext. Data Fig.7 | miR-29c-3p | CTQ0 vs CTQ2 | Mann-Whitney test | 21/18 | U = 154 | 0.3351 | CTQ0: 0.76 [0.53–1.33]; CTQ2: 0.6 [0.38–1.3] |
| Ext. Data Fig.7 | miR-148a-3p | CTQ0 vs CTQ2 | Mann-Whitney test | 19/19 | U = 177 | 0.9310 | CTQ0: 0.94 [0.47–1.57]; CTQ2: 0.89 [0.48–1.9] |
| Ext. Data Fig.7 | miR-101-3p | CTQ0 vs CTQ2 | Mann-Whitney test | 21/20 | U = 167 | 0.2709 | CTQ0: 0.86 [0.43–1.37]; CTQ2: 0.42 [0.25–1.4] |
| Ext. Data Fig.8 | miR-145-5p | Cell line, type of serum and interaction | Two-way ANOVA + Tukey’s test | 6–9/ group | Interaction: F = 1.79×10^6^; DF = 1  Cell line: F = 1.79×10^6^; DF = 1  Serum: F = 1.66; DF = 1 | Interaction: 0.9989; Cell line: 0.9989; Serum: 0.2083 | Tukey: NS vs SRB1 KD (all comparisons p ≥ 0.7872) |
| Ext. Data Fig.9 | miR-223-3p | Control vs MSUS (serum) | Unpaired t-test | 6/12 | t = 1.01; df = 16.0 | 0.3255 | Control: 1 ± 0.3; MSUS: 1.16 ± 0.31 |
| Ext. Data Fig.9 | miR-145-5p | Control vs MSUS (serum) | Mann-Whitney test | 6/12 | U = 35 | 0.9461 | Control: 1.03 [0.81–1.16]; MSUS: 1.04 [0.89–1.11] |
| Suppl. Fig.1 | Significant anxiety symptoms | Control vs PLMS (children) | Fisher’s exact test | 35/70 | OR = 1.55 | 0.4924 | Control: 27/8; PLMS: 48/22 |
| Suppl. Fig.1 | Generalized anxiety disorder | Control vs PLMS (children) | Fisher’s exact test | 35/70 | OR = 1.8 | 0.3338 | Control: 29/6; PLMS: 51/19 |
| Suppl. Fig.1 | Panic disorder | Control vs PLMS (children) | Fisher’s exact test | 35/70 | OR = 1.55 | 0.4924 | Control: 27/8; PLMS 48/22 |
| Suppl. Fig.3 | Total reads | Control vs PLMS (children) | Unpaired t-test | 18/48 | t = 1.83; df = 64 | 0.0727 | Control: 1.8×10^7^ ± 5.3×10^6^; PLMS: 2.1×10^7^ ± 5.8×10^6^ |
| Suppl. Fig.3 | Too short reads | Control vs PLMS (children) | Unpaired t-test | 18/48 | t = 2.02; df = 64 | **0.048** | Control: 8.5×10^6^ ± 2.9×10^6^; PLMS: 1.0×10^7^ ± 3.6×10^6^ |
| Suppl. Fig.3 | miRNAs | Control vs PLMS (children) | Mann-Whitney test | 18/48 | U = 400 | 0.653 | Control: 4.0×10^6^ [1.7×10^6^–7.3×10^6^]; PLMS: 4.8×10^6^ [1.9×10^6^–8.2×10^6^] |
| Suppl. Fig.3 | piRNAs | Control vs PLMS (children) | Mann-Whitney test | 18/48 | U = 410 | 0.759 | Control: 2.0×10^5^ [9.0×10^4^–3.9×10^5^]; PLMS: 1.6×10^5^ [8.8×10^4^–3.0×10^5^] |
| Suppl. Fig.3 | Other RNA species | Control vs PLMS (children) | Mann-Whitney test | 18/48 | U = 416 | 0.825 | Control: 1.0×10^6^ [5.8×10^5^–1.7×10^6^]; PLMS: 1.2×10^6^ [6.5×10^5^–1.5×10^6^] |
| Suppl. Fig.3 | Other | Control vs PLMS (children) | Mann-Whitney test | 18/48 | U = 275 | **0.0233** | Control: 3.0×10^6^ [2.5×10^6^–3.4×10^6^]; PLMS: 3.6×10^6^ [3.0×10^6^–4.3×10^6^] |
| Suppl. Fig.5 | Trauma severity | CTQ2 | Ordinary one-way ANOVA | 8–23/ group | F = 0.78; DF = 5.92 | 0.7825 |  |
| Suppl. Fig.7 | Total reads | CTQ0 vs CTQ2 | Welch’s t-test | 26/23 | t = 0.07; df = 36.29 | 0.9408 | CTQ0: 3.2×10^7^ ± 5.8×10^6^; CTQ2: 3.2×10^7^ ± 9.2×10^6^ |
| Suppl. Fig.7 | Too short reads | CTQ0 vs CTQ2 | Mann-Whitney test | 26/23 | U = 295 | 0.9446 | CTQ0: 1.7×10^7^ [1.3×10^7^–2.0×10^7^]; CTQ2: 1.7×10^7^ [1.2×10^7^–2.1×10^7^] |
| Suppl. Fig.7 | miRNAs | CTQ0 vs CTQ2 | Mann-Whitney test | 26/23 | U = 288 | 0.8349 | CTQ0: 3.1×10^6^ [2.2×10^6^–3.8×10^6^]; CTQ2: 2.9×10^6^ [1.8×10^6^–4.1×10^6^] |
| Suppl. Fig.7 | piRNAs | CTQ0 vs CTQ2 | Mann-Whitney test | 26/23 | U = 292 | 0.8973 | CTQ0: 3.3×10^5^ [2.0×10^5^–4.5×10^5^]; CTQ2: 3.5×10^5^ [1.9×10^5^–4.8×10^5^] |
| Suppl. Fig.7 | Other RNA species | CTQ0 vs CTQ2 | Unpaired t-test | 26/23 | t = 0.0006; df = 47 | 0.9995 | CTQ0: 5.4×10^6^ ± 2.6×10^6^; CTQ2: 5.4×10^6^ ± 2.9×10^6^ |
| Suppl. Fig.7 | Other | CTQ0 vs CTQ2 | Mann-Whitney test | 26/23 | U = 270 | 0.5713 | CTQ0: 3.1×10^6^ [2.1×10^6^–6.6×10^6^]; CTQ2: 3.6×10^6^ [2.4×10^6^–6.3×10^6^] |
| Suppl. Fig.8 | miR-34c | Number of CT events vs VST expression | Spearman correlation | 66 | r = -0.07 | 0.55 |  |
| Suppl. Fig.8 | miR-449-5p | Number of CT events vs VST expression | Spearman correlation | 66 | r = -0.02 | 0.88 |  |
| Suppl. Fig.9 | WHO criteria | CTQ0 vs CTQ2 | Fisher’s exact test | 40/28 | OR = 1.17 | 0.8071 | CTQ0: 23/17; CTQ2: 15/13 |
| Suppl. Fig.10 | miR-320e | Control vs PLMS (adults) | Mann-Whitney test | 14/9 | U = 27.5 | **0.0240** | Control: 0.71 [0.58–1.49]; PLMS: 5.08 [0.67–8.57] |
| Suppl. Fig.11 | SRB1 | NS vs SRB1 KD | Unpaired t-test | 3/3 | t = 18.08; df = 4 | **<0.0001** | NS: 1 ± 0.06; SRB1 KD: 0.28 ± 0.03 |
| Suppl. Fig.18 | miR-223-3p | Serum vs sperm miRNA expression | Pearson correlation | 11 | r = 0.53, 95% CI [-0.11, 0.86]; df = 9 | 0.0953 |  |
| Suppl. Fig.18 | miR-145-5p | Serum vs sperm miRNA expression | Pearson correlation | 11 | r = 0.34, 95% CI [-0.33, 0.78]; df = 9 | 0.3088 |  |
